# A Comparison of In-Person and Telehealth Treatment Modalities using the SpeechVive Device

**DOI:** 10.1101/2025.01.16.25320611

**Authors:** Renee Covert, Sandy Snyder, Ashleigh Lambert, Mary Spremulli, Brianna Blandford, Kaitlyn Dwenger, Georgia Malandraki, Meghan McDonough, Françoise Brosseau-Lapre, Jessica E. Huber

**Affiliations:** Department of Speech, Language, and Hearing Research, Purdue University, West Lafayette, IN 47907; HCA Healthone Presbyterian/St. Luke’s, Denver, CO 80218; Voice Aerobics, LLC, Punta Gorda, FL; Faculty of Kinesiology, University of Calgary, Calgary, Alberta; Department of Communicative Disorders and Sciences, University at Buffalo, Buffalo, NY 14214

## Abstract

Telehealth is increasing popular as a treatment option for people with Parkinson disease (PD). The SpeechVive device is a wearable device that uses the Lombard effect to help patients speak more loudly, slowly, and clearly. This study sought to examine the effectiveness of the device to improve communication in people with PD, delivered over a telehealth modality as compared to in-person, using implementation science design. 66 people with PD were enrolled for 12 weeks with 34 choosing the in-person group and 32 in the telehealth group. Participants were assessed pre-, mid-, and post-treatment. Participants produced continuous speech samples on and off the device at each timepoint. Sound pressure level (SPL), utterance length, pause frequency, and total pause duration were measured. Psychosocial surveys were administered to evaluate the effects of treatment on depression, self-efficacy, and participation. The in-person group increased SPL when wearing the device while the telehealth group did not. Both groups paused less often while wearing the device. Utterance length increased post-treatment for the telehealth group, but not for the in-person group. An increase in communication participation ratings in the telehealth group, but not the in-person group, was the only significant change in the psychosocial metrics. The in-person group showed similar treatment effects as previous studies. The device was not as effective in the telehealth group. One limitation was data loss due to recording issues that impacted the telehealth group more than the in-person group.

## INTRODUCTION

Parkinson disease (PD) is the second most common neurodegenerative disease of adulthood following Alzheimer’s disease (Aarsland et al., 2007). PD causes an aggregation of alpha synuclein proteins throughout the brain, predominantly in the basal ganglia. This results in impairments across multiple systems, including cognitive, motor, and sensory systems responsible for speech. The most common speech impairment is hypokinetic dysarthria, which is characterized by hypophonia, variable rate, breathiness, monopitch and monoloudness, and imprecise consonants. Hypophonia is a major contributor to decreased intelligibility in parkinsonian speech (Ho et al., 1999). Increased loudness, attention, and speech effort are major foci of speech therapy for those with PD.

The predominant goal in speech therapy for people with PD is to increase internally perceived effort to improve overall loudness and intelligibility. There are multiple therapies that are used to target hypokinetic dysarthria in people with PD. One common therapy is the Lee Silverman Voice Treatment (LSVT). This protocol has been developed to emphasize high-effort loud phonation over the course of four one-hour sessions for four weeks to improve vocal intensity (SPL) and intelligibility (Pu et al., 2021; L. O. Ramig et al., 2001). Another treatment used to improve the loudness and intelligibility for people with PD is the SpeakOut!®, often combined with group therapy, but less research has been conducted on this program. SpeakOut!® is based on the hypothesis that ongoing maintenance is required to retain therapeutic gains due to the progressive nature of PD (Behrman et al., 2020). The SpeakOut® program is a standardized program of twelve individual, forty-minute sessions over the course of four weeks. Patients are encouraged to begin attending a group-oriented maintenance-based therapy during the third or fourth week of SpeakOut®. The program’s focus is on speaking loudly and intentionally. SpeakOut® has also been shown to be effective in improving loudness, intonation, and voice quality (Behrman et al., 2020; Levitt et al., 2015).

The SpeechVive device is a newer technique that differs from the other two behavioral therapies. This is a device-driven treatment focusing on improving loudness, rate, and articulatory clarity. This device triggers the Lombard effect in speakers by playing multi-talker babble into one ear as they speak. The Lombard effect is a reflex that causes a speaker to talk more loudly, clearly, and slowly in the presence of background noise (Pick et al., 1989). The Lombard effect has been found to be intact in people with PD (Adams & Lang, 1992; Stathopoulos et al., 2014a). SpeechVive has been shown to increase loudness both immediately (Stathopoulos et al., 2014a) and after 8-weeks using the device (Richardson, Huber, Kiefer, Kane, et al., 2022; Richardson, Huber, Kiefer, & Snyder, 2022). After 8 weeks of using the device, people with PD increased utterance length and decreased rate of speech (Richardson, Huber, Kiefer, & Snyder, 2022, 2022). In addition, individuals using the SpeechVive rated the cognitive demands of using the device as less demanding when compared to the LSVT LOUD treatment (Richardson, Huber, Kiefer, & Snyder, 2022). While the SpeechVive device has been shown to be efficacious in in-person and highly controlled environments (randomized control trials), there is insufficient information regarding its efficacy in telehealth modalities or as implemented in clinical practice.

During the COVID-19 pandemic, there was an increase in the utilization of and need for telehealth services across the healthcare field. In addition, the COVID-19 pandemic highlighted disparities in accessing telerehabilitation services due to internet access and reliability of signal, technology training and comfortability, and reimbursement issues limiting access to high quality treatment and care for many patients, especially those in rural areas (Canoro et al., 2023; Hassan et al., 2020; Hirko et al., 2020). Even with the pandemic increasing the number of articles examining telehealth in a variety of healthcare fields, there is still a significant lack of research around telehealth in speech therapy for individuals with PD.

There is some research to suggest that speech teletherapy with people with PD is effective and preferred (Constantinescu et al., 2010; Dias et al., 2016; Theodoros et al., 2006a). The Lee Silverman Treatment Program (LSVT) has shown success when delivered through telehealth. Theodoros et al. (2006) performed a treatment study to investigate the treatment of PD over a 4-week period using LSVT via a telehealth platform. This study recruited 10 participants who completed teletherapy via the eREHAB system from another room at the clinic on a lab-provided computer. They found that there were significant improvements in breathiness, loudness, pitch, and loudness variability after treatment. Based on participant survey data, they found overall satisfaction with the treatment method.

A later article by Constantinescu et al. (2011) compared the effectiveness of LSVT delivered in-person vs. telehealth in a non-inferiority trial. This study included 32 participants. Both groups received therapy at the University of Queensland with the telehealth group being in a different room on a different floor from the therapist. They found significant changes in sound pressure level (SPL) without significant difference between the groups. Their survey data found the online group to be both happy and comfortable in the online session. The study also did not report any major technological difficulties with adequate video and audio quality. However, they noted some difficulty with network congestion as well as microphone and speech processing malfunction.

A more recent article by Griffin et al. (2017) further looked at the efficacy of LSVT in a non-inferiority trial. This study included 29 participants, 8 in the telehealth group. Telehealth participants received LSVT treatment via an iPad within their home. Both groups showed improvement in SPL from baseline.

An additional article by Theodoros et al. (2016) evaluated the effectiveness of telehealth treatment utilizing the LSVT treatment approach in a non-inferiority randomized control trial. This study included participants living within and outside the metropolitan area. The 32 metropolitan participants were randomized into in-person and telehealth groups, while 21 rural participants were allocated into a rural telehealth group. Did not find group differences in SPL but did find statistically significant improvements in SPL measures. In addition, they reported changes in the dysarthria impact profile but no changes in the Parkinson’s Disease Questionnaire-39 (PDQ-39).

These studies suggest that people with PD are not only successful with telehealth/homebased treatment methods but prefer it. While these studies are promising, more research into the effectiveness of teletherapy with people PD is needed to fully understand how best to implement this delivery paradigm in the field of speech-language pathology. Previous studies were limited by a small sample size, internet connectivity issues, and the fact that participants were not in their own home. While some studies (Griffin et al., 2017) have found similar results when providing treatment inside the patient’s home, they were still limited by many of the other variables including internet connectivity and small sample size. It is of vital importance to test the efficacy of telehealth in a comfortable and real-life environment for people with PD. Technological literacy, internet access, and caregiver interaction need to be assessed as supporting or confounding variables. In addition, many technological advances have been made recently which may improve or diminish the relevance of these results, necessitating more up-to-date research. While these advances in in-home testing and more modern technology are beginning to be studied, there is minimal data regarding speech treatment outcomes for people with PD. Further, it is not clear if the results for telehealth hold for all treatment techniques since the speech telehealth studies have only examined LSVT.

Other fields have also investigated the efficacy of telehealth treatment and rehabilitation for people with PD, and have shown that it is highly effective (Cikajlo et al., 2018; Colón-Semenza et al., 2018; Durner et al., 2017; Quinn et al., 2020). For instance, Colón-Semenza et al. (2018) performed a feasibility study analyzing the efficacy of physical therapy (PT) treatments and peer coaching delivered through an online modality and reported improvement in mobility and a high satisfaction rate amongst study participants. Outcomes for gait, balance, and medical management were similar for teletherapy as in-person sessions. Gandolfi et al. (2017) performed a study analyzing the efficacy of an in-home PT treatment based on the Nintendo Wii Fit system compared to another treatment performed in clinic. They found that the online modality was a feasible alternative when used for reducing postural instability (Gandolfi et al., 2017).

Telemedicine has also been rated more preferred by those with PD in comparison to in-person treatment, especially for medical treatment (Beck et al., 2017a; Dobbs et al., 2018; Durner et al., 2017; Wilkinson et al., 2016). Beck et al. (2017a) examined the provision of neurological care through in-person care augmented with telehealth visits compared to usual in-person care to those with PD. They found that treatment outcomes did not differ between the groups. Additionally, they found that 97% of the patients and 86% of the providers were satisfied or very satisfied with the telemedicine treatment modality. Dorsey et al. (2016) had a similar design when testing PT delivered over an online modality. Their findings were similar to Beck et al. (2017b), but they also found that technological difficulties could be a major barrier.

While many of these studies across disciplines have shown that teletherapy is effective, very few have done so in “real life” clinical conditions. Implementation-based science is a vital element of clinical research, examining whether results from randomized control studies can be demonstrated with real life variables in a typical clinical environment. These variables can include varying treatment goals and administration approaches, insurance and administrative barriers related to duration and frequency of sessions, computer connectivity difficulties, patient preferences and mindset, and human variability.

PD has been shown to not only cause physical changes as a result of the systems affected, but patients often experience psychological changes in concordance with their diagnosis. These changes can include increased depression, anxiety, and apathy, among others (Broen et al., 2016; Hanna & Cronin-Golomb, 2012; Kirsch-Darrow et al., 2011; Pedersen et al., 2009; Pluck, 2002). These psychological changes can have a negative impact on a patient’s quality of life (Duncan et al., 2014; Hanna & Cronin-Golomb, 2012). Studies evaluating the impact of PD related communication changes on quality of life found decreased quality of life scores and communication participation cited as a result of PD (Miller et al., 2006; Takahashi et al., 2016).

Only a handful of studies have evaluated the impact of speech therapy on communication-based quality of life measures. One study by Ramig et al. (2018) found participant rated communicative effectiveness (CETI-M) to significantly increase with LSVT treatment. A meta-analysis evaluating LSVT across 10 studies, 4 used the Voice Handicap Index to evaluate voice-related quality of life changes (H. Xu et al., 2020). All 4 found VHI changes after LSVT treatment.

Psychological changes can impact therapy effectiveness and should be considered during assessment and treatment of speech impairments. If treatment results in improvements in psychosocial outcomes, this could signal better health-related quality of life and generalized functionality which are the goals of rehabilitative therapy. Psychosocial changes have predominantly been measured using the PDQ-39 in both medical and rehabilitation disciplines. Some studies have found overall score decreases and therefore increases in reported quality of life in PDQ-39 scores pre and post treatment in both medicine and speech therapy disciplines (Beck et al., 2017c; Theodoros et al., 2016). However, these studies found variability between the sub-sections and the changes were not statistically significant between group or time differences. In addition, none of these studies used validated measures of these psychosocial constructs. This makes it difficult to draw conclusions regarding the efficacy of intervention on quality of life related outcomes for those with PD.

More research is needed examining treatment-related psychosocial changes utilizing surveys validated to evaluate these specific measures. The Geriatric Depression Scale (GDS) has been validated and tested to screen for depression in older adults and people with PD. Beck et al. (2017b) used this scale to evaluate changes in depression when administering remote neurologic care to patients with PD. However, they did not find any significant changes in depression ratings after the medical treatment. Another study, utilizing balance training to improve gait and balance related difficulties associated with PD, reported improvements in GDS scores for patients after receiving the treatment (Smania et al., 2010). However, it is unclear if the effects were from treatment or from exercise-related effects which has been shown to improve depression independently (Kim et al., 2023). These mixed results indicate that different treatment modalities and disciplines may have different effects on depression in PD warranting further research.

Another common symptom that can impact treatment outcomes is apathy. However, few studies have evaluated how therapy outcomes can impact apathy ratings. The Starkstein Apathy Scale, validated for people with PD, has been used to evaluate treatment-related changes in apathy in studies particularly focusing on increasing physical activity. These studies have found conflicting results. While some have shown decreased apathy scores (Cugusi et al., 2015), others have found unchanged apathy scores (Inoue et al., 2021). Another study utilizing virtual music therapy targeting apathy for people with PD reported an improvement in apathy scores measured by the Starkstein Apathy Scale (Shah-Zamora et al., 2024). While this scale can be used to show treatment-related changes in apathy, it is unclear if these changes are due to increased social or physical activity or if the results would also be seen in other disciplines like speech therapy.

The Neuro-QOL Satisfaction with Social Roles and Activities-Short Form and Ability to Participate in Social Roles and Activities-Short Form are two surveys that have been developed and validated to evaluate the impact of neurodegenerative illness on quality of life, including those with PD. However, we have been unable to identify studies that evaluated treatment-related changes in these outcomes in people with PD. This is a vital area of research to evaluate how therapies can motivate bigger changes in participation and satisfaction.

The Communication Participation Item Bank (CPIB) is a validated measure to evaluate communicative participation in a variety of populations. We have been unable to find studies that report treatment-related changes in the CPIB. However, a study by Theorodos et al (2016) utilized the Dysarthria Impairment Profile to evaluate changes in speech-related quality of life. This study found significant reduction in psychosocial impact of the participant’s speech disorder as well as improved acceptance of their dysarthria with no difference between in-person and telehealth groups after LSVT. They did not find significant changes across the profile sub-sections evaluating the effect of the dysarthria on the person, the way participants communicated following treatment, or others’ reaction to the speech disorder. It is of vital importance that the field of speech therapy not only understands the acoustic and perceptual changes that can be achieved in therapy, but also the impact of speech therapy on psychosocial outcomes that can improve patient-reported communicative quality of life. However, the relationship between speech therapy for people with PD and patient-reported communicative quality of life is not yet fully understood.

Within the realm of teletherapy for those with PD, a handful of studies have reported slight improvements in quality of life, quality of communication, and satisfaction when delivered via telehealth as compared to in-person treatment (Sekimoto et al., 2019; Y. Xu et al., 2022). Xu et al. (2022) surveyed 944 patients with PD regarding their opinions on telehealth. Participants reported the highest rates of telehealth satisfaction with speech therapy services (52/66) and mental health related appointments (95/137). Some reasons for increased satisfaction included “reduced travel time and ease and convenience” (p. 1290). Some participants indicated that telehealth can be limiting for more complicated appointments and had a higher preference for in-person appointments, particularly those involving physical exams. However, other studies have not found a difference in quality of life ratings when using telehealth modalities compared to in-person modalities (Beck et al., 2017c). There is a clear need for additional research into the impact of treatment on the psychosocial outcomes for people with PD, particularly when the treatment is provided via teletherapy.

### Purpose

The first aim of this study was to evaluate the efficacy of the SpeechVive device in increasing sound pressure level (SPL), while decreasing pause frequency and duration in an implementation study design, using the in-person group data only. Some studies have shown that the device is effective in these areas; however, this research has been predominantly in randomized control studies. The second aim of this study was to evaluate the effectiveness of the SpeechVive device when administered over teletherapy, comparing the in-person and telehealth group data. The third aim of this study is to evaluate if treatment with the SpeechVive device improved patient reported psychosocial and quality of life outcome measures in both the in-person and telehealth group data.

## METHODS

### Participants□□

This study was approved by the Purdue University Institutional Review Board (#1602017214).

One hundred and thirteen (113) people with PD and their caregivers were screened for participation in this study. For inclusion, patients were required to have a diagnosis of PD by a neurologist and a diagnosis of hypokinetic dysarthria from a speech-language pathologist; be stimulable to the Lombard effect; be willing to participate in therapy; have internet access with adequate bandwidth, webcam access, and an email address that is checked regularly; be a native speaker of English; and have a caregiver who was willing to participate. Caregivers were required to score in the normal range on the Montreal Cognitive Assessment (Nasreddine et al., 2005). Exclusionary criteria included a history of neurological diseases other than PD and use of bilateral hearing aids (since one ear must be free for the SpeechVive device). Seventeen screened participants were excluded and 30 decided not to participate for a variety of reasons (**Figure 1**).

**Figure 1.**
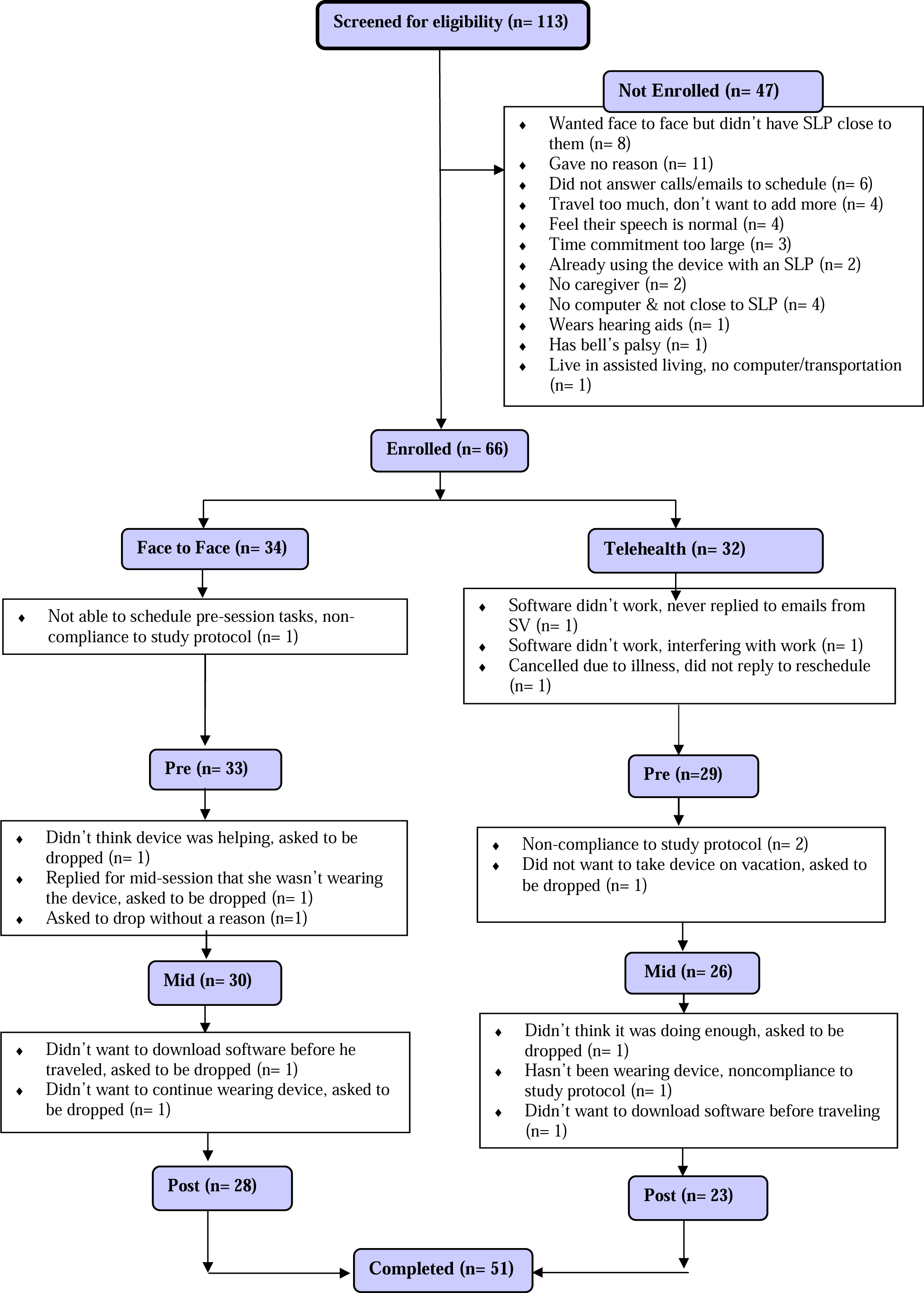
Participant CONSORT.

A total of 66 participants were included in the study. Participants chose if they preferred to participate in the telehealth (32 participants, 13 female) or in-person groups (34 participants, 11 female). A total of 23 telehealth and 28 in-person participants completed the study (

**Table *1***).□In cases where participants dropped out after the pre-treatment session, the last assessment point was analyzed in alignment with intent-to-treat principles. The mean age of the telehealth group was 60 years (SD=14.4 years) and 13 participants had deep brain stimulation (DBS) implants. The mean age of the in-person group was 67 years (SD=19.3 years), and 8 participants had DBS implants. The ages between groups was statistically significant. Both groups were living in the community and had a moderate range of motor severity and moderate speech severity. Participants lived in a range of communities with a bias toward suburban areas. All but 10 participants were on a form of carbidopa levodopa (Sinemet) or dopamine agonist. Full participant demographics can be found in **Table *1***, individual participant data can be found in **Table B10**. Caregivers were primarily spouses of the participants with PD.

**Table 1.** Participant Demographics.

|  | In-person | Telehealth | Entire Study |
| --- | --- | --- | --- |
| Females, Males | 11, 23 | 13,18 | 24, 41 |
| Race | 74% Caucasian<br>6% Black/African American<br>6% American Indian/Alaskan Native<br>14% Did not disclose | 94% Caucasian<br>3% Two or More Races<br>3% Asian<br>3% Did not disclose | 80% Caucasian<br>3% American Indian/Alaskan Native<br>3% Black/African American<br>2% Asian<br>2% Two or More Races<br>11% Did not disclose |
| Ethnicity | 82% Not Hispanic/Latino<br>18% Did not disclose | 97% Not Hispanic/Latino<br>3% Did not disclose | 88% Not Hispanic/Latino<br>11% Did not disclose |
| Mean Age (SD) | 67 (19.3) | 60 (14.4) | 66.2 (10.1) |
| Age Range | 44-88 | 37-78 | 37-88 |
| Mean Speech Severity (SD) | 42.7 (20.8) | 43.6 (22.3) | 43.1 (21.3) |
| Mean Time Since Dx | 10.6 (8) | 10.6 (8.7) | 10.6 (8.3) |
| On carbidopa/levodopa and/or dopamine agonist | 91% | 90% | 91% |
| Region | Rural 16%<br>Urban 16%<br>Suburban 68% | Rural 33%<br>Urban 14%<br>Suburban 52% | Rural 25%<br>Urban 15%<br>Suburban 58% |

*Note: 27% of participants did not provide information about the region where they lived*

### Therapy□□

After completing informed consent, participants with PD were matched with a clinically certified speech-language pathologist (SLP) in their state who was participating in the research study. Clinicians only provided treatment in states where they were licensed and for the treatment modality for which they could bill. Of the 11 SLPs involved in the study, 8 clinicians provided only in-person treatment while 3 provided both telehealth and in-person treatment. All participants were treated and assessed during their medication ‘on’ state.

For the SpeechVive condition, participants were asked to wear the device 2-8 hours a day during times that they were communicating. They were also asked to read aloud with the device for 30 minutes a day 5 days a week. The SLPs were responsible for programming the SpeechVive device. This included obtaining fundamental frequency to develop the window for determining voicing onset and obtaining SPL without the device. The device sensitivity was then adjusted to ensure the noise the device emitted was consistently turning on and off with the participant’s voice. During reading or conversational speech, the intensity of the noise from the device was set to elicit a 3-5 dB SPL increase over the participant’s comfortable intensity. Programming and device adjustment were performed via the SpeechVive interface with the device connected to the computer through the USB port.

The SLP developed and administered treatment plans according to goals set by the clinician and patient as an element of the implementation design. All patients used the SpeechVive device, but some participants also received other therapy during the study period, consistent with SLP and patient goals (**Figure 2**). Treatment differences between groups (in-peron and telehealth) were minimal and most treatment was provided in a group setting. No participants received dysphagia treatment throughout the duration of this study. For the telehealth group, three participants received additional therapy sessions. One participant attended two group sessions associated with SpeakOut!®. One person received a 30-minute treatment session on rate and pausing. One person received a one hour and 15-minute session on rate and clear speech. For the in-person group, 17 participants received additional therapy sessions. Most of these sessions were group treatment associated with SpeakOut!; 10 participants received an average of 4.8 hour-long sessions (range 1-10). Lastly, four participants received an average of 2.25 treatment sessions focused on rate, pausing, and voice of approximately 30-minutes each (range 1-4).

**Figure 2.**
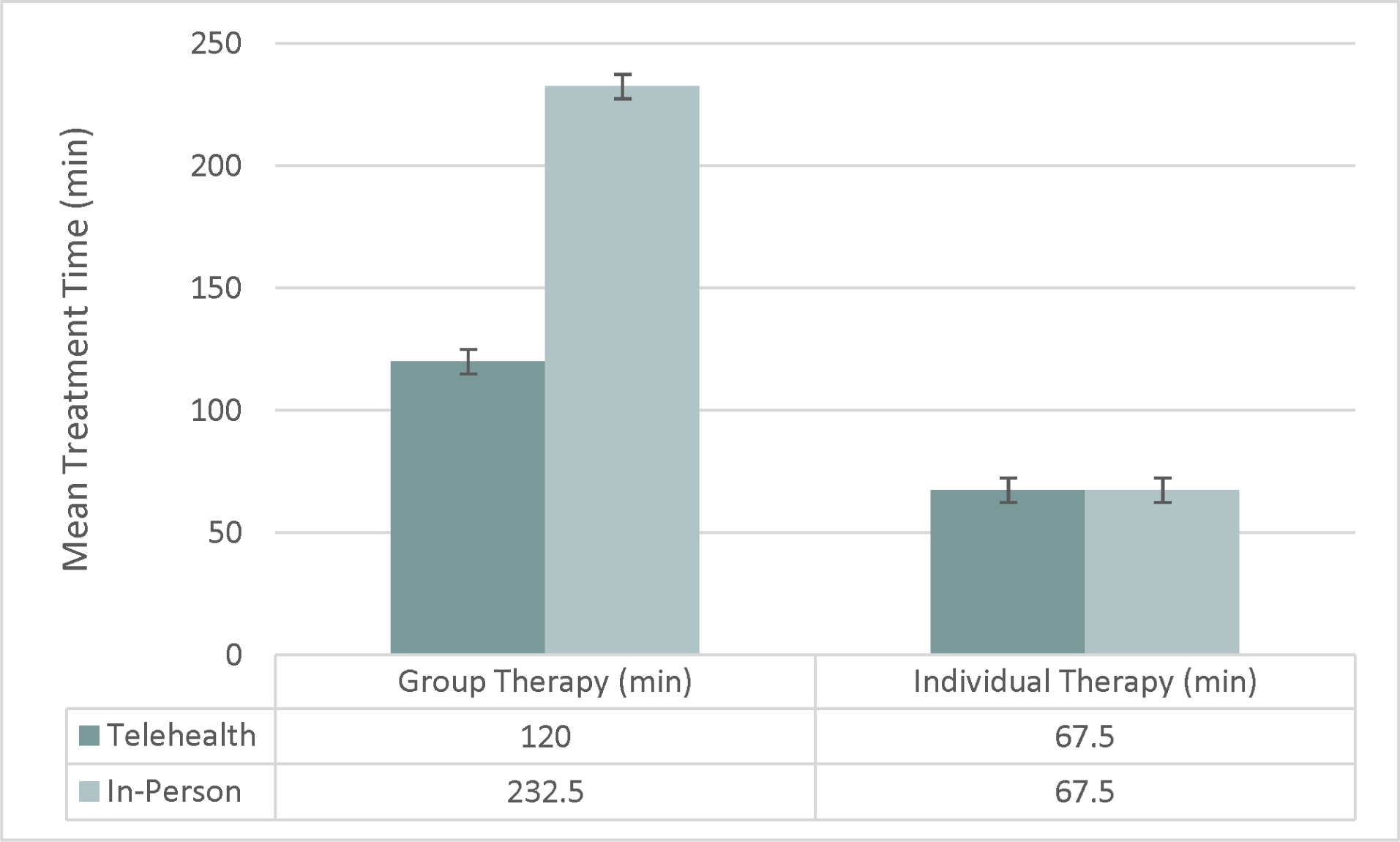
Mean Supplemental Treatment Time.

### Data Collection

Data was collected in assessment sessions before treatment (pre), after 6 weeks of treatment (mid), and after 12 weeks of treatment (post) in the same modality that treatment was administered (in-person or telehealth). Midpoint data was only used for participants when the post-treatment data was unavailable due to drop-out. Data was collected by the SLPs who were trained in data collection, and audio recordings were obtained through software developed to interface with the SpeechVive device and record audio data. A table-top microphone (Samson Go) was used to collect data directly into the SpeechVive software at 24 kHz through the USB port. The mouth-to-microphone was held constant at 12 inches.

#### Speech Tasks

Participants completed a 2-minute monologue on a topic of their choice (how they met their spouse, about their children or pets, a book or movie they enjoyed) as well as a reading of the Rainbow Passage first without wearing the device (off condition) and then again with the device on (on condition). The recordings were then uploaded to FileLocker by the SLP (in-person) or the patient (telehealth).

The samples were orthographically transcribed by an undergraduate research staff member and checked by a second undergraduate research staff member. Difficult passages were discussed among research staff to support valid transcriptions.

#### Acoustic Outcome Measurements

**Vocal intensity**, as measured by sound pressure level (SPL in dB), was the primary outcome measure for this study. Vocal intensity is perceptually correlated with perceived loudness, a common deficit in hypokinetic dysarthria. Mean SPL for each utterance was obtained through a Praat script (Boersma & Weenink, 2008). Utterances were delimited by pauses of 150 ms or greater. Secondary outcomes were chosen based on reported changes in speech of people with PD in prior literature. People with PD have been reported to produce shorter utterances, pause more frequently and for longer duration, and speak at a faster rate of speech than age-matched control speakers (Huber & Darling, 2011). **Utterance length** was determined by counting the number of syllables produced per utterance. **Articulation rate** (in syllables per second) was calculated by dividing the number of syllables in an utterance by the utterance duration. **Pause frequency and duration** (in seconds) were measured as well. All measurers were blinded to the group (telehealth or in-person) and device condition (off or on the device) to support unbiased results.

Due to internet connectivity issues and difficulty with data collection, there were some issues with the microphone signal (loss of signal, repeating signal, overlapping speech). Protocols were developed to reliably identify and remove microphone signal issues to not compromise SPL measures. Overlapping speech occurred when the SLP, who was collecting data, talked over the participant. There were also some issues with background noise in the SLP’s office or the home where the participant was located during the recordings that caused data loss. In cases where there was noise or overlapping speech, duration was obtained from the utterances and pauses, and if the syllables were clearly identifiable in Praat, utterance length was calculated. However, SPL was not measured from utterances where the microphone signal was noisy or when overlapping speech occurred. When internet connectivity caused the loss of signal or a signal that skipped or repeated, we measured what was available. If the signal was lost, we did not measure any of the outcomes for the utterance. In some cases, we could obtain durations of pauses or utterances and utterance lengths reliably in these files. In all cases, we tried to maintain as much data as possible. For example, utterances were parsed to isolate the areas of recording issues or overlapping speech to allow the measurement of SPL from those parts of the utterance that were unaffected by the recording issues (**Figure 3**). Syllable and duration measures were taken from the entire utterance if they were preserved. If these issues were present for greater than or equal to 75% of the sound file, the task was not measured. **Table *2***. Vocal Intensity Data Missing by Session**Table 2** provides data on the sessions where no SPL data was available. This reflects the worst case for missing data since for many of those sessions, utterance length, and utterance and pause duration information were available.

**Figure 3.**
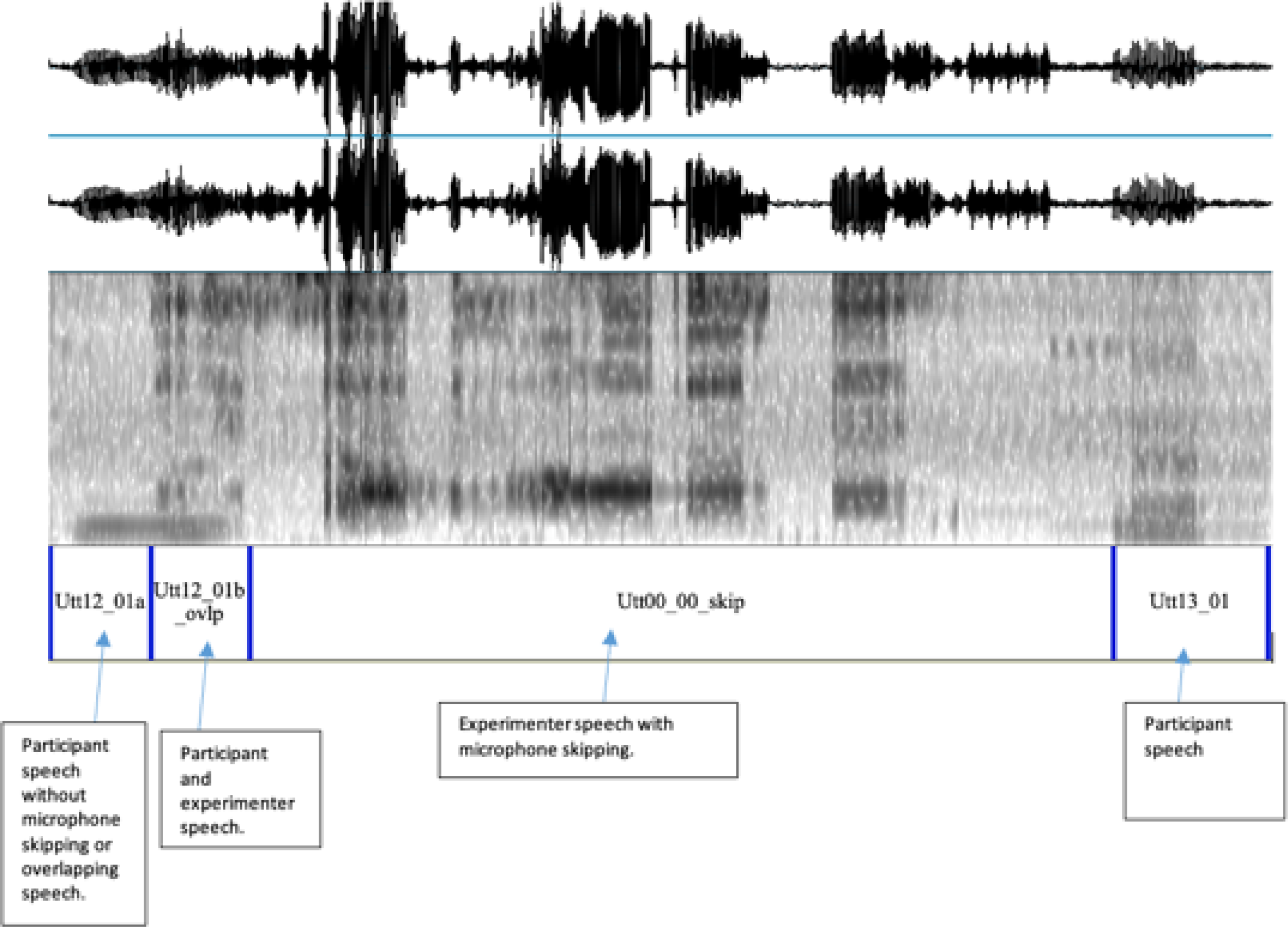
Speech sample with microphone skipping and overlapping speech.

**Table 2.**
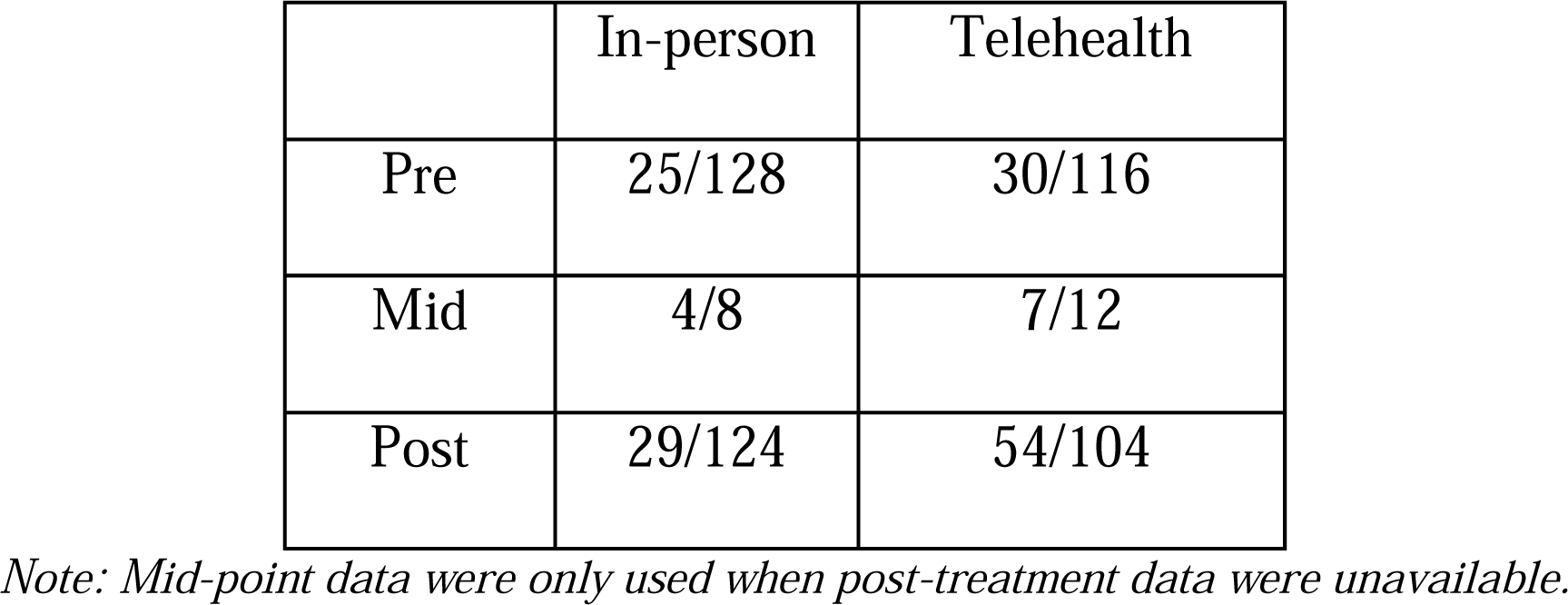
Vocal Intensity Data Missing by Session.

### Questionnaires□

Pre-treatment all participants completed a health questionnaire developed in the laboratory that asked about medications, timing of medications, PD symptoms, voice symptoms, and other health conditions. Medication information was checked at mid-treatment data collection sessions. Pre- and post-treatment all participants completed questionnaires evaluating psychosocial factors including quality of life, depression, anxiety, communication participation, independence in activities of daily living, impact of PD on life, and relationship satisfaction. Post-treatment all participants completed a SpeechVive satisfaction survey and a time and travel survey. The telehealth group additionally completed a tele-satisfaction survey adapted from Sharma et al., 2013. Replacement of missing data was addressed by mean substitution using the mean response for the questions that were answered at the same time point from the same participant.

The Geriatric Depression Scale, Short Form (Yesavage & Sheikh, 1986) was used to assess levels of depression. The scale has been standardized on individuals 55 years and older and has been found to effectively assess levels of depression in people with PD (Mantri et al., 2022; Schrag et al., 2007). This questionnaire contained 15 items, and participants were asked to assign yes/no answers to each item based on how they had felt over the last week. Higher scores indicate higher levels of depression, with a score higher than 5 suggestive of depression.

Starkstein Apathy Scale (Starkstein et al., 1992) was utilized to assess the participant’s perception of their apathy symptoms. This scale has been validated and used to assess levels of apathy within people with PD. It contains 14 items scored on a scale from 0 (not a lot) to 3 (a lot), with higher scores indicating less apathy.

The Neuro-QOL Satisfaction with Social Roles and Activities-Short Form and Ability to Participate in Social Roles and Activities-Short Form (Cella et al., 2012) were used to assess perceived ability and satisfaction with social interactions. These measures were developed to assess quality of life across neurologic conditions assessing physical, social, and mental health and standardized on participants with neurological conditions including ALS, CVA, epilepsy, MS, and PD. They have been validated for use with people with PD in additional studies (Nowinski et al., 2016). It contains 8 items scored on a scale from 1 to 5, with 5 indicating more satisfaction and participation in social interactions.

The Communication Participation Item Bank, Short Form (Baylor et al., 2013) was used to assess participant perception of communication participation. This questionnaire contains 10 speaking conditions like communicating in a small group or getting a turn in a fast-moving conversation. We asked participants how their PD impacted their ability to participate in these communication conditions. Scores for each item ranged from 0 (very much) to 3 (not at all), with higher scores indicating higher levels of communication participation. Scores were converted to T-scores, per the guidelines in Baylor et al. (2013). If mean substitution resulted in a score that included decimals, the score was rounded before the T-score conversion.

A time and travel survey was used to assess the perceived cost and benefit of telehealth and in-person care in relation to access, ability, and willingness to travel (see Appendix A.1). The survey was developed by the researchers for this study. It consisted of 10 multiple choice, yes/no, and open response questions.

A SpeechVive satisfaction survey was developed by the researchers to assess overall satisfaction with the SpeechVive device across both groups (see Appendix A.2). The survey consisted of 6 questions asking the participants to rate on a scale of 1 (strongly disagree) to 5 (strongly agree) with higher scores indicating higher levels of satisfaction.

A telehealth satisfaction survey was adapted from Sharma et. al (2013). This survey was used to assess overall satisfaction with the telehealth treatment modality (see Appendix A.3). It asked participants to rate their agreement with 14 statements about telehealth from 1 (strongly disagree) to 5 (strongly agree) with higher scores indicating higher levels of satisfaction.

### Statistics and Reliability

Inter-measurer reliability was completed for 20% of the pre- and post-sessions. A different research assistant than the person who completed the original measures completed the second measurement. Inter-rater reliability was quantified using interclass correlations (two-way mixed, single measurement, absolute agreement) and judged using guidelines published by Koo and Li (2016)

To assess changes in acoustic outcome measurements, we used a mixed model analysis of variance (ANOVA) with a between factor of group (telehealth vs. in-person) and two within factors of time (pre vs. post) and device condition (device off vs. device on). Participant and treating SLP were included as random factors. Data was entered at the utterance level. Tukey honestly significantly different tests were used to assess any significant interactions of group by time (session). Cohen’s D was used to assess the effect sizes of any significant interactions. Alpha level was set as p<.05.

To assess changes in patient-related outcome measures, we used a mixed model analysis of variance (ANOVA) with a between group factor of group (telehealth vs. in-person) and a within factor of time (pre vs. post). Participant and treating SLP were included as random factors. Tukey honestly significantly different tests were used to assess any significant interactions of group by time. For the SpeechVive satisfaction scale, a t-test was used to compare the telehealth group to the in-person group. Missing data were handled by mean substitution using the mean response for the questions that were answered at the same time point for the same participant. Cohen’s D was used to assess the effect sizes of any significant interactions. Alpha level was set as p<.05.

## RESULTS

### Aims Reliability

Interclass correlations (ICC) for the inter-rater reliability reflected good (.75 to.9) to excellent (>.9) reliability (Koo & Li, 2016). For SPL, the ICC was .837 [95% confidence interval (CI): .828-.846]. For utterance length in syllables, the ICC was .774 [95% CI: .762-.785]. For utterance duration in seconds, the ICC was .796 [95% CI: .785-.806]. For the number of pauses, the ICC was .925 [95% CI: .885-.951]. For pause durations in seconds, the ICC was .957 [95% CI: .930-.973].

### Speech Production Outcomes (Aims 1 and 2)

Results from the ANOVAs are presented in **Table 3**. Alpha-levels for pairwise comparisons are presented in text, with effect sizes for significant findings. Means and standard deviations are presented by group, device condition, and time for each outcome in **Table *4***.

**Table 3:** Statistical Outcomes for Speech Measures.

|  | Group<br>(tele/in-person) |  | Time (pre/post) |  | Group by<br>Time |  | Device<br>Condition<br>(on/off) |  | Device<br>Condition by<br>Group |  | Time by<br>Device<br>Condition |  | Time by<br>Device<br>Condition by<br>Group |  |
| --- | --- | --- | --- | --- | --- | --- | --- | --- | --- | --- | --- | --- | --- | --- |
|  | F<br>value | P<br>value | F<br>value | P<br>value | F<br>value | P<br>value | F<br>value | P<br>value | F<br>value | P<br>value | F<br>value | P<br>value | F<br>value | P<br>value |
| **SPL** | 3.01 | 0.0827 | **150.46** | **<.0001** | **19.12** | **<.0001** | **143.39** | **<.0001** | **119.42** | **<.0001** | 0.29 | 0.5904 | 3.74 | 0.0531 |
| **Utterance**<br>**Length** | 3.72 | 0.0539 | 0.32 | 0.5687 | **6.39** | **0.0115** | **12.69** | **0.0004** | 0.96 | 0.3281 | 2.29 | 0.1299 | 0 | 0.9923 |
| **Utterance**<br>**Duration** | **11.84** | **0.0006** | 2.06 | 0.1512 | 3.6 | 0.0579 | **13.54** | **0.0002** | 0.48 | 0.4873 | 1.73 | 0.1883 | 0.05 | 0.8177 |
| **Articulation**<br>**Rate** | 0.23 | 0.6323 | **17.70** | **<.0001** | 0.56 | 0.4542 | 3.71 | 0.0541 | 2.29 | 0.1300 | 2.15 | 0.1423 | 0.81 | 0.3685 |
| **Total**<br>**number of**<br>**pauses** | 0.06 | 0.8049 | 0.7 | 0.4028 | 0.16 | 0.6854 | **6.74** | **0.0099** | 0.04 | 0.8374 | 0.21 | 0.6461 | 0 | 0.9562 |
| **Total Pause**<br>**Duration (s)** | 1.31 | 0.2537 | 0.01 | 0.9315 | 0.07 | 0.7939 | **6.28** | **0.0127** | 0.43 | 0.5133 | 0.45 | 0.5031 | 0.02 | 0.8993 |
| **Average**<br>**Duration (s)** | 1.08 | 0.2296 | 0 | 0.9711 | 0.48 | 0.4893 | 0.27 | 0.6059 | 0 | 0.9843 | 0.75 | 0.3872 | 0.62 | 0.4311 |

**Table 4.** Means and Standard Deviation for Speech Production Measures.

|  | Device<br>Setting | Pre |  | Post |  |
| --- | --- | --- | --- | --- | --- |
|  |  | In-Person | Tele | In-Person | Tele |
| **Sound Pressure**<br>**Level (SPL)** | OFF | 69.87 (6.89) | 72.40 (5.51) | 68.20 (8.25) | 71.86 (7.29) |
|  | ON | 71.92 (6.60) | 73.09 (6.22) | 71.12 (7.48) | 72.43 (7.97) |
| **Utterance Length**<br>**(syllables)** | OFF | 8.14 (6.26) | 9.51 (7.06) | 7.97 (5.99) | 9.89 (7.68) |
|  | ON | 8.39 (6.17) | 9.87 (6.81) | 8.38 (6.24) | 10.43 (7.51) |
| **Utterance**<br>**Duration (sec)** | OFF | 1.91 (1.31) | 2.27 (1.59) | 1.85 (1.20) | 2.28 (1.59) |
|  | ON | 1.98 (1.26) | 2.38 (1.54) | 1.92 (1.24) | 2.41 (1.58) |
| **Articulation Rate**<br>**(syll/sec)** | OFF | 4.16 (1.22) | 4.17 (1.33) | 4.17 (1.23) | 4.30 (1.52) |
|  | ON | 4.12 (1.22) | 4.18 (1.35) | 4.27 (1.28) | 4.31 (1.51) |
| **Total Number of**<br>**Pauses** | OFF | 43.4 (17.59) | 40.67 (10.95) | 40.87 (15.87) | 39.09 (13.09) |
|  | ON | 39.77 (18) | 36.82 (12.94) | 37.89 (14.88) | 36.89 (11.74) |
| **Total Duration of**<br>**Pauses (sec)** | OFF | 37.32 (16.74) | 30.8 (9.67) | 36.15 (19.34) | 30.38 (10.02) |
|  | ON | 33.45 (15.83) | 27.97 (11.78) | 32.61 (17.2) | 29.09 (13) |
| **Average Duration**<br>**of Pauses (sec)** | OFF | 1.01 (1.3) | .80 (.27) | .87 (.34) | .82 (.25) |
|  | ON | .92 (.52) | .77 (.24) | .89 (.40) | .79 (.20) |

### SPL (dB)

Results revealed significant time, group by time, device condition, and group by device condition effects in SPL (**Table *3***). No other main or interaction effects were significant. For the group by time effect (**Figure 4**), participants in both the telehealth and in-person groups significantly decreased their mean SPL after treatment (**Table *4***). The telehealth group mean SPL decreased by .61 dB (p<.001, d_RM_ = -.072 [-.417 to .311]), and the in-person group mean SPL decreased 1.27 dB (p<.001, d_RM_ = -.099 [-.437 to .311]). There was no significant difference between the groups at either pre- (p=.467) or post-treatment (p=.183). For the group by device condition effect (**Figure *5***), participants in the in-person group increased their mean SPL by 2.45 dB when the device was turned on (p<.0001, d_RM_ = -.282 [ -.595 to .156]). Participants in the telehealth group increased their mean SPL by .67 dB when the device was turned on (p<.0001 d_RM_ = .069 [-.330 to .417]). There was no significant difference between the groups for the device on (p=.714) or device off (p=.076) conditions.

**Figure 4.**
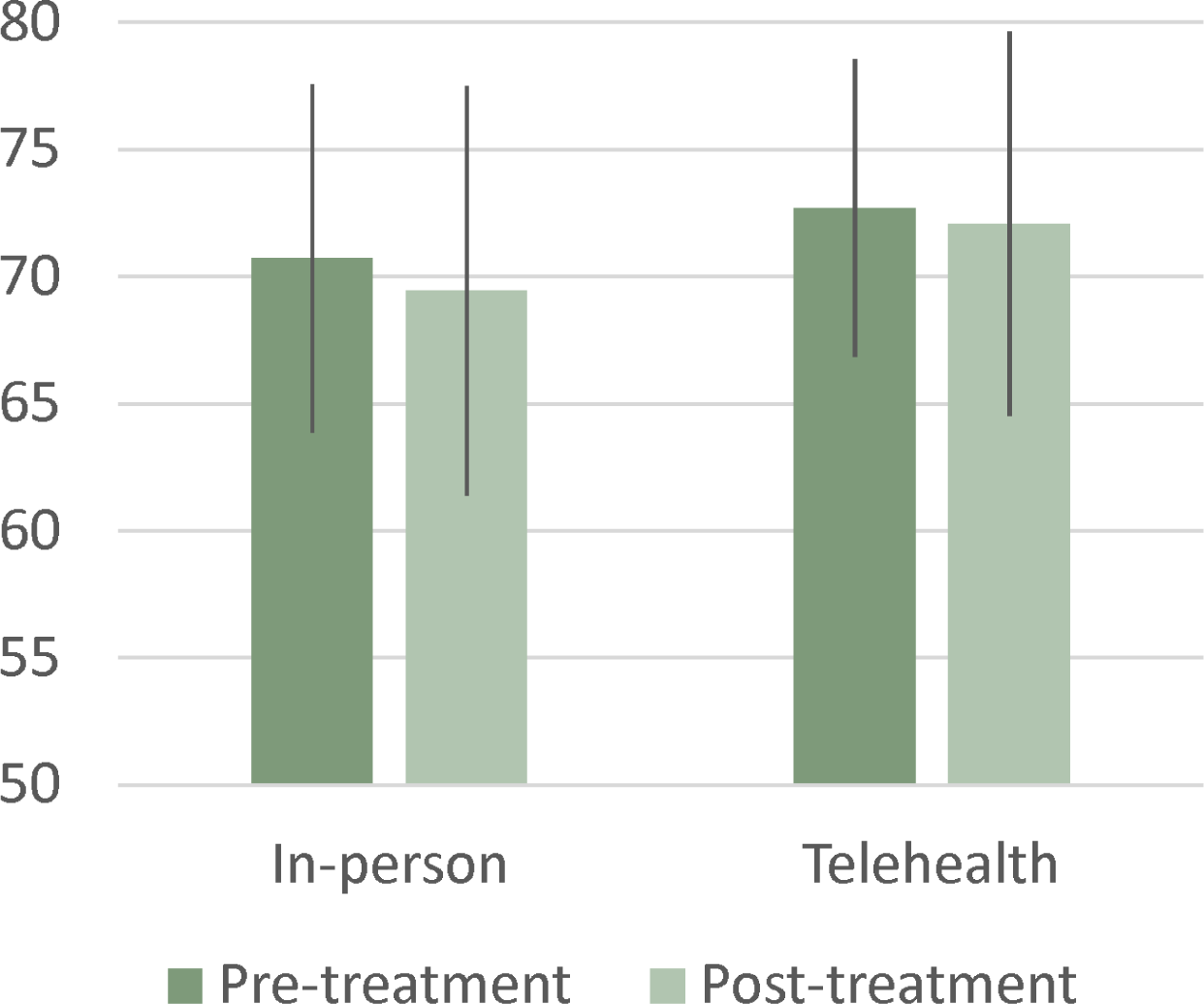
SPL Group by Time Effects (p<.001).

**Figure 5.**
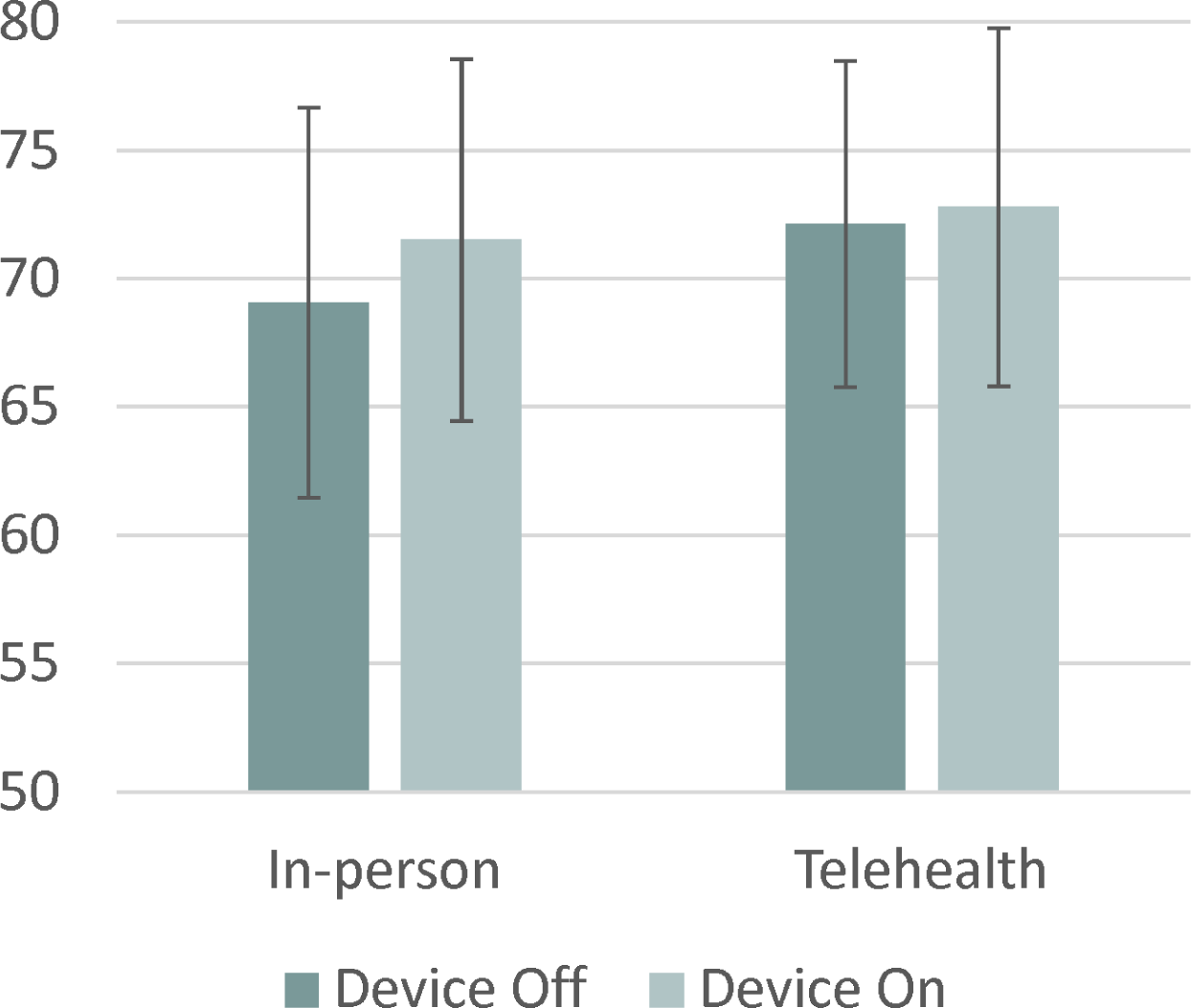
SPL Group by Device Condition Effects (p<.001.)

### Utterance Length (syllables)

Results showed significant group by time and device condition effects in utterance length. There was an increase in utterance length between pre- and post-treatment in the telehealth group (+.43 syllables) and a decrease in utterance length in the in-person group (-.10 syllables). However, neither of these were significant in pairwise comparisons after Tukey-Kramer adjustment (telehealth p=.19, d_RM_ = .044 [ -.344 to .403]) (in-person p=.40, d_RM_ = -.011 [-.382 to .365]). For the device condition effect, participants increased utterance length by .37 syllables (p=.0004, d_RM_ = .038 [-.347 to .401]).

### Utterance Duration (sec)

Results revealed significant group and device condition effects in utterance duration. For the group effect, participants in the telehealth group had a longer utterance duration than those in the in-person group (p= .0006, d_RM_ = 237 [-.226 to .524]). For the main effect of device condition, participants increased their utterance duration with the device turned on as compared to the device turned off (p=.0002 d_RM_ = .042 [-.345 to .403]).

### Articulation Rate (syllables per second)

Results indicated a significant time effect for rate of speech. Participants increased their articulation rate from pre- to post-treatment by about .09 syllables per second (d_RM_ = .052 [-.337 to .410]).

### Number of Pauses

Results showed a significant device condition effect for number of pauses. There was a mean decrease in the number of pauses with the device turned on (M = -3.3 pauses or about 7.8% fewer pauses) (d_RM_ = -.034 [-.48 to 2.69]).

### Pause Duration (sec)

Results revealed a significant device condition effect for the total pause duration across the sample, but not for average pause duration. On average, the total duration of pauses decreased when the device was on (M = -3.12 sec) (d_RM_ = -.138 [-.469 to .279]).

### Patient Reported Outcome Measure Outcomes (Aim 3)

Statistical results for each survey questionnaire are presented in **Table 5**. Means and standard deviations are presented by group and time for each questionnaire in **Table *6***.

**Table 5.** Statistical Results for Survey Data.

|  | Group |  | Time |  | Group by Time |  |
| --- | --- | --- | --- | --- | --- | --- |
|  | F value | P value | F value | P value | F value | P value |
| GDS | .87 | .3550 | 1.41 | .2410 | .71 | .4046 |
| Apathy | .51 | .4773 | 3.7 | .0607 | .43 | .5164 |
| CPIB | 3.37 | .0728 | 1.35 | .2505 | **5.15** | **.0280** |
| Neuro A | 3.83 | .0564 | .05 | .8233 | 1.04 | .3135 |
| Neuro B | .34 | .5640 | .16 | .6935 | .67 | .4175 |

**Table 6.** Means with Standard Deviations for Survey Questionnaires.

|  | Pre |  | Post |  |
| --- | --- | --- | --- | --- |
|  | In-Person | Tele | In-Person | Tele |
| GDS | 3.09 (3.11) | 2.52 (2.23) | 3.16 (3.48) | 2.53 (2.13) |
| Apathy | 11.35<br>(5.08) | 9.26 (6.01) | 12.12<br>(6.44) | 10.73 (5.6) |
| CPIB | 47.38<br>(8.53) | 40.57<br>(10.06) | 45.92<br>(9.88) | 45.07<br>(11.16) |
| Neuro A | 31.02<br>(5.81) | 28.82<br>(6.24) | 30.29<br>(7.34) | 30.46<br>(6.32) |
| Neuro B | 27.16<br>(7.66) | 25.62<br>(7.56) | 26.34<br>(7.86) | 27.06<br>(7.51) |

### Psychosocial Surveys

There were no significant differences between the two treatment groups for any survey outcomes (**Table *5***). Pre and post survey data is visualized in **Figure *7***

**Figure 6.**
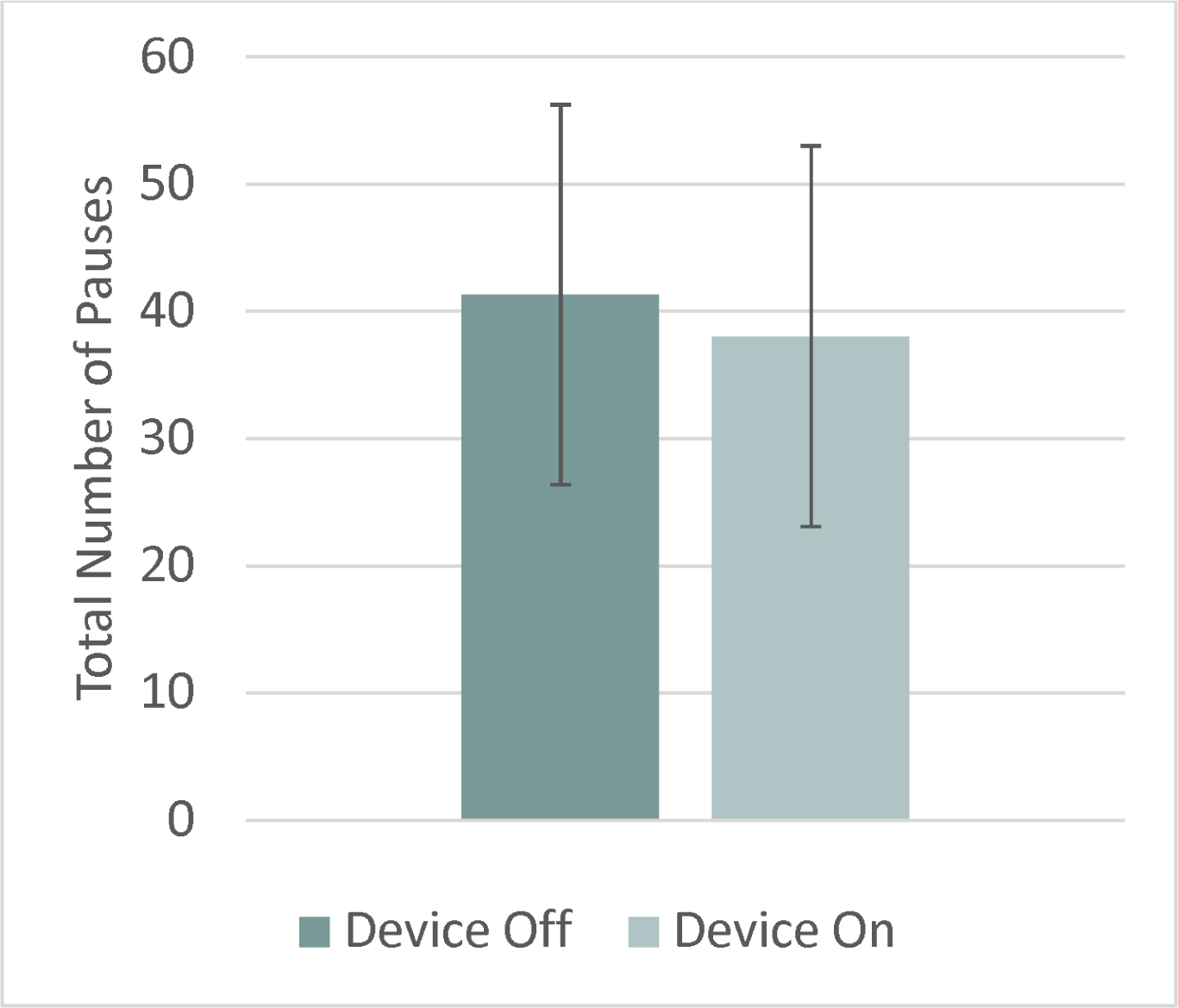
Number of Pauses Device Condition Effect (p=.0099).

**Figure 7.**
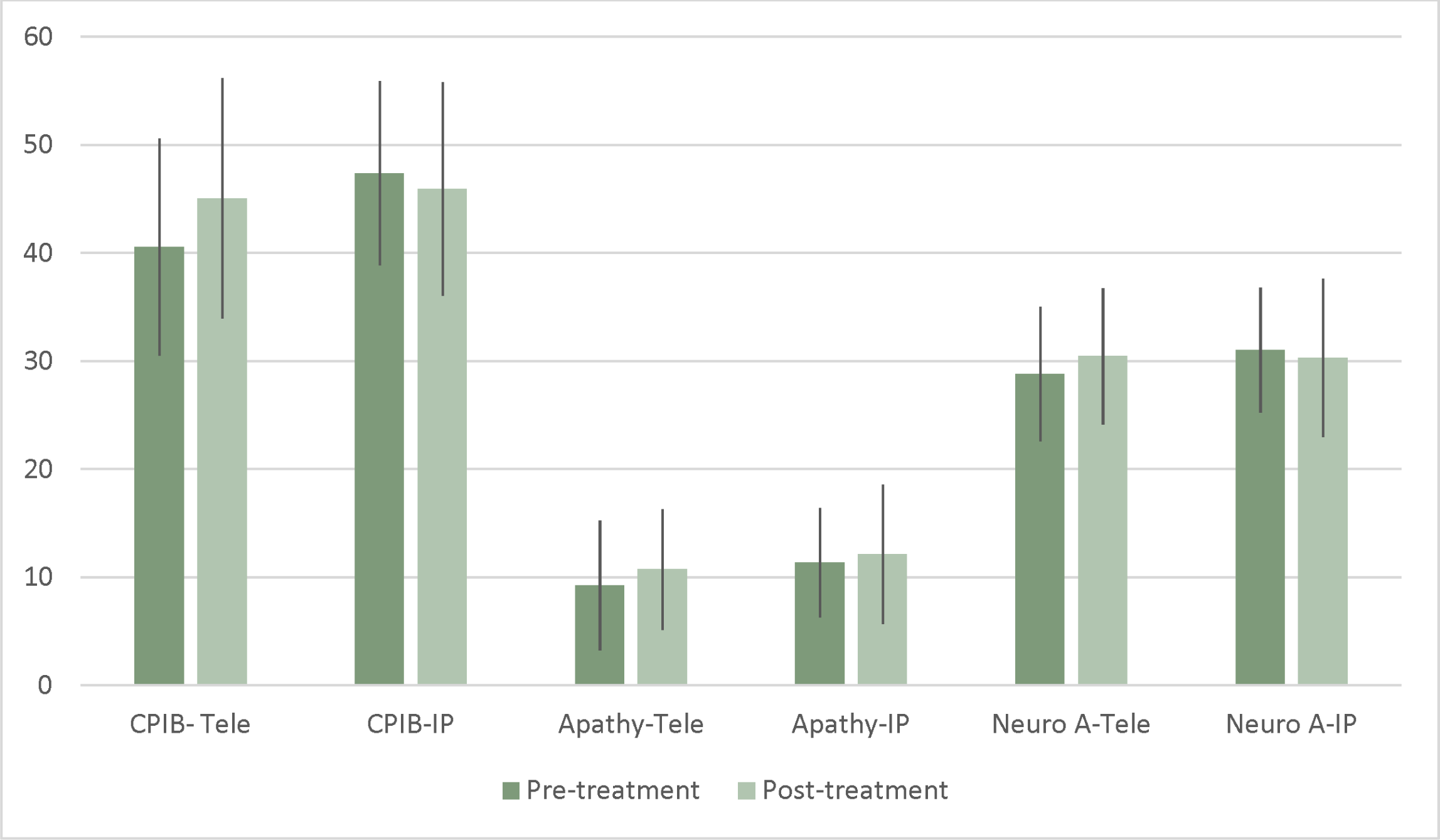
Pre-Post Survey Data.

The CPIB showed a significant group by time effect; however, effect was not significant in pairwise comparisons after Tukey-Kramer adjustment was applied. There were no significant differences from pre- to post-treatment for the in-person group (p=.85, d_RM_ = -.118 [.517-.205]) or the telehealth group (p=.09, d_RM_ = .41 [.141-.87]). However, the effect size for the telehealth group was large. Ratings increased more in telehealth group after treatment (+4 z-score points) than in-person (-1 z-score points). There were no significant group differences at pre-treatment (p=.05) or post-treatment (p=.87).

Analysis did not indicate significant effects of time, group, or group by time in the GDS, Starkstein Apathy Scale, or the Neuro-QOL Ability to Participate in Social Roles and Activities-Short Form (Neuro B). Some of the changes were close to significant. We highlight these here, given the small sample size for survey research. Apathy scale scores increased more for participants in the telehealth group (+1.47) than the in-person group (+.77) after treatment (Table 6). Neuro-QOL Satisfaction with Social Roles and Activities-Short Form (Neuro A) scores increased more for the telehealth group (+1.64) than the in-person group (-.73).

### SpeechVive Satisfaction Survey

T-tests did not demonstrate a significant difference between groups (t= -.49, p=.63) regarding satisfaction with the SpeechVive device. Out of a total score of 30, with higher scores reflecting greater satisfaction, the mean response for the in-person group was 24.11(SD=4.38), and the mean response for the telehealth group was 23.64 (SD=3.92). Score distribution across participants is shown in **Figure *8***.

**Figure 8.**
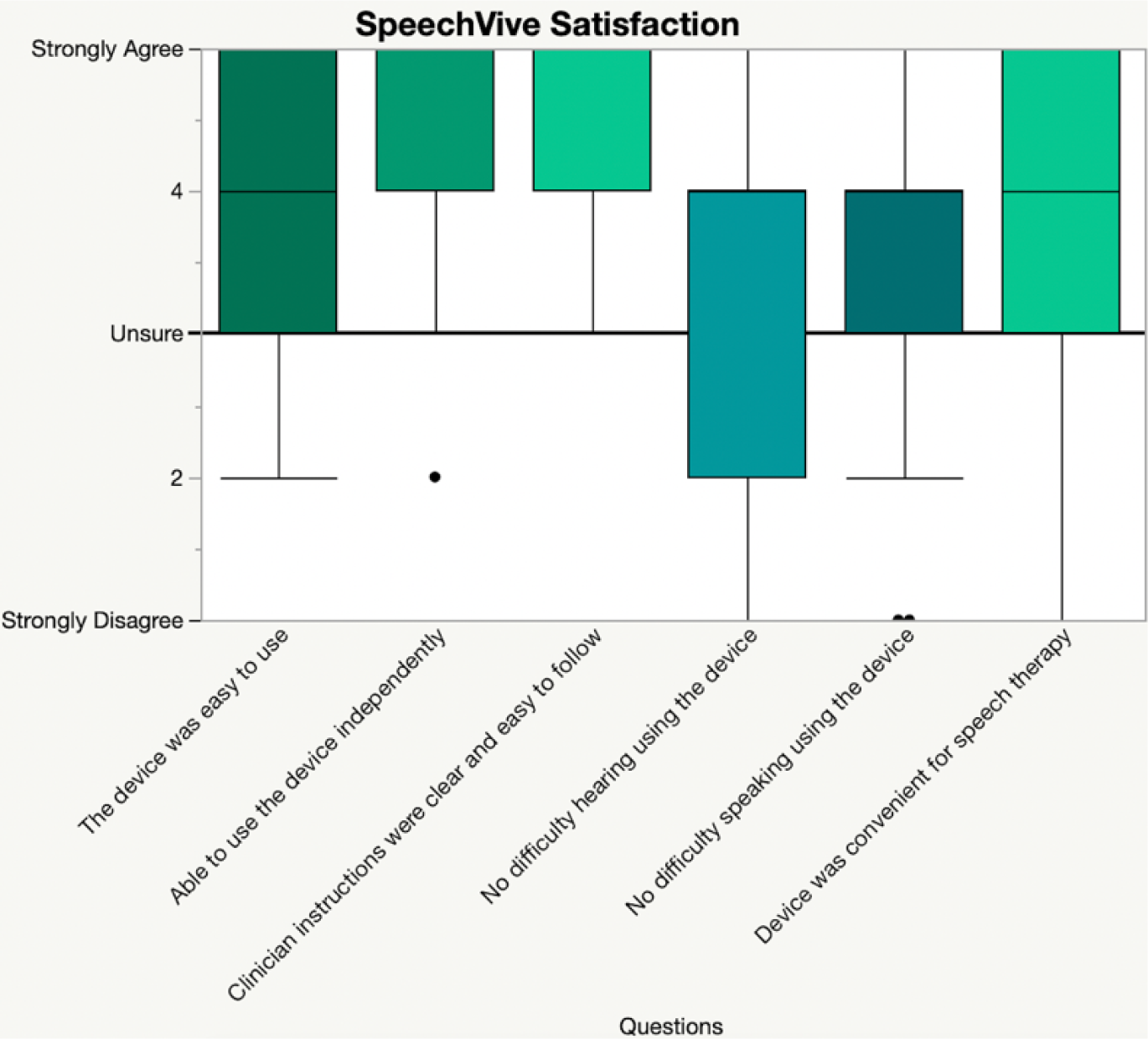
SpeechVive Satisfaction Box and Whisker.

### Telehealth Satisfaction Survey

Participants in the telehealth group had an average satisfaction of 59.1 (SD=10.61) out of a total score of 70, with higher scores indicating higher levels of satisfaction with the telehealth services. The highest score was a 69 while the lowest satisfaction score was a 29. Survey ratings across participants is demonstrated in **Error! Reference source not found.**.

### Time and Travel Survey

In the telehealth group, 11 participants lived in a suburban setting, 8 lived in a rural setting, and 3 lived in an urban setting. In the in-person group, 7 participants lived rurally, 17 lived in a suburban setting, and 2 lived in an urban setting.

There was a wide range in responses for both the telehealth and in-person groups as to where the nearest speech therapist lives (see Table 7). It is unclear in some responses if the participant was indicating the location of the nearest accessible speech therapist, or the location of the speech therapist in the study. For the telehealth group, responses ranged from incredibly close (0-10 miles) to incredibly far (over 100). There was less variation in the in-person group with most having access to an SLP within 10 miles and most not going beyond 50.

**Table 7.** Distance to Nearest Speech Therapist.

|  | In-person | Teletherapy |
| --- | --- | --- |
| 0-10 miles<br>(including virtual) | 11 | 8 |
| 11-20 miles | 3 | 0 |
| 21-30 miles | 2 | 3 |
| 31-40 miles | 4 | 1 |
| 41-50 miles | 3 | 0 |
| 51-60 miles | 0 | 0 |
| 61-70 miles | 0 | 1 |
| 71-80 miles | 1 | 0 |
| 81-90 miles | 0 | 0 |
| 100+ miles | 0 | 3 |

Participants in the telehealth group indicated that 41% of them had a working caregiver while only 35% of people in the in-person group had a working caregiver. In the telehealth group 32% of participants indicated they travel alone to therapy and 27% indicated they travel alone in general. On the other hand, 42% of the in-person group indicated they travel alone to therapy and 35% indicated they travel alone in general.

There was significant variation within the amount of time it took for people to travel/prepare for a therapy session in both groups. In the in-person group, there was a wider variety in time taken, with participants noting it took anywhere between less than ten minutes to over 60 minutes to prepare for a session. 43% of participants reported that it took 20 minutes or less to prepare for the session. On the other hand, it didn’t take longer than 40 minutes for anyone in the telehealth group, with 76% of participants reporting 20 minutes or less for preparation time. There was also significant variation in the amount of time it took to transition back into normal activities after a session, which includes travel time. In the in-person group, subjects reported it taking less than ten minutes to over 60 minutes to transition back to normal activities. The greatest amount of people (50%) reported it took less than ten minutes to transition back, while 20% of people reported that it took greater than 30 minutes to transition back. In the telehealth group, participants reported the time ranging between 0-20 minutes to transition back to normal activities. The greatest amount of people (81%) indicated it taking less than 10 minutes while the remaining participants reported it taking no more than 20 minutes to return to normal activities. The time taken to prepare for and transition away from therapy is shown in **Table *7*** and **Figure *10*** and **Error! Reference source not found.**.

**Figure 9.**
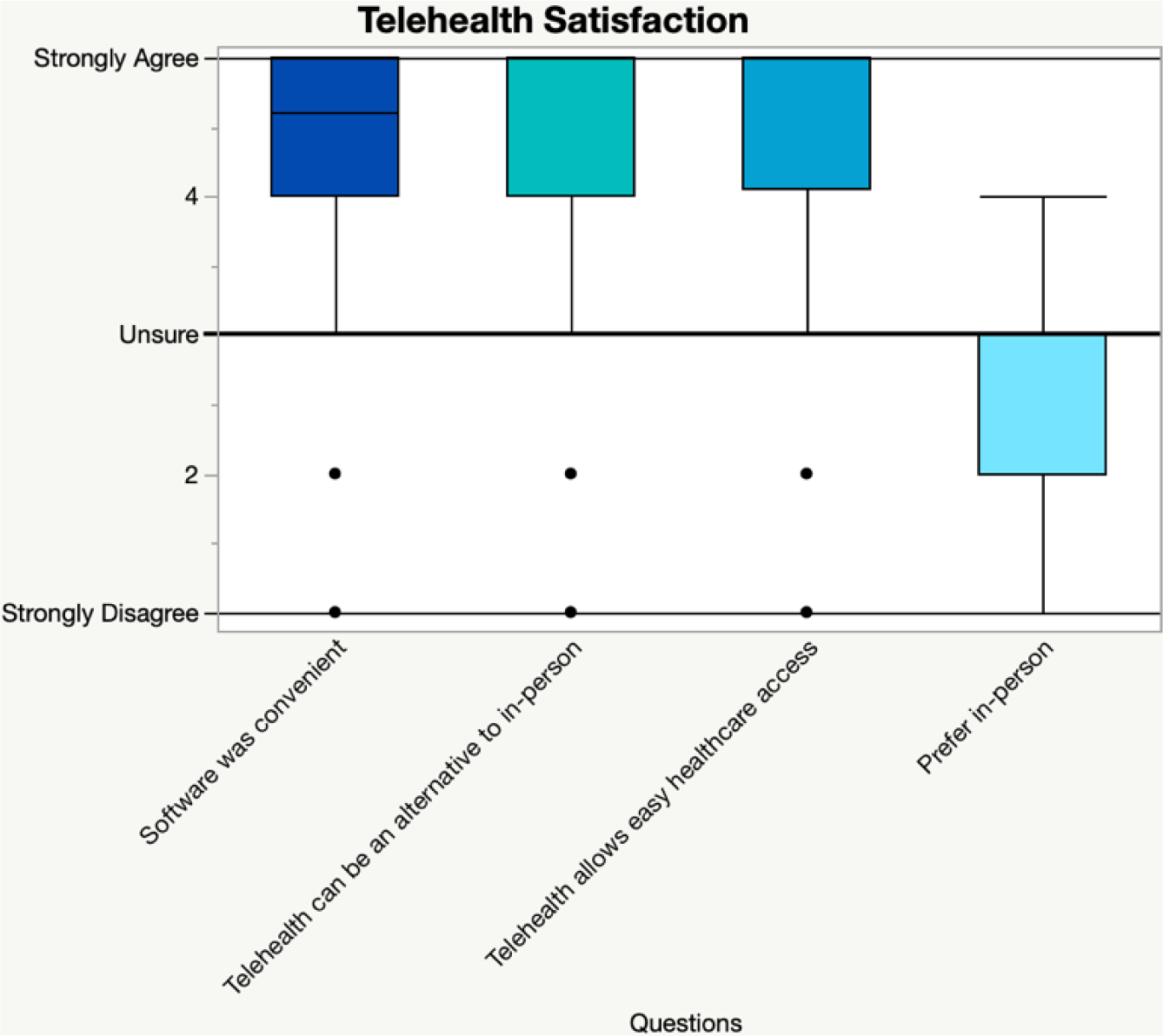
Telehealth Satisfaction Box and Whisker.

**Figure 10.**
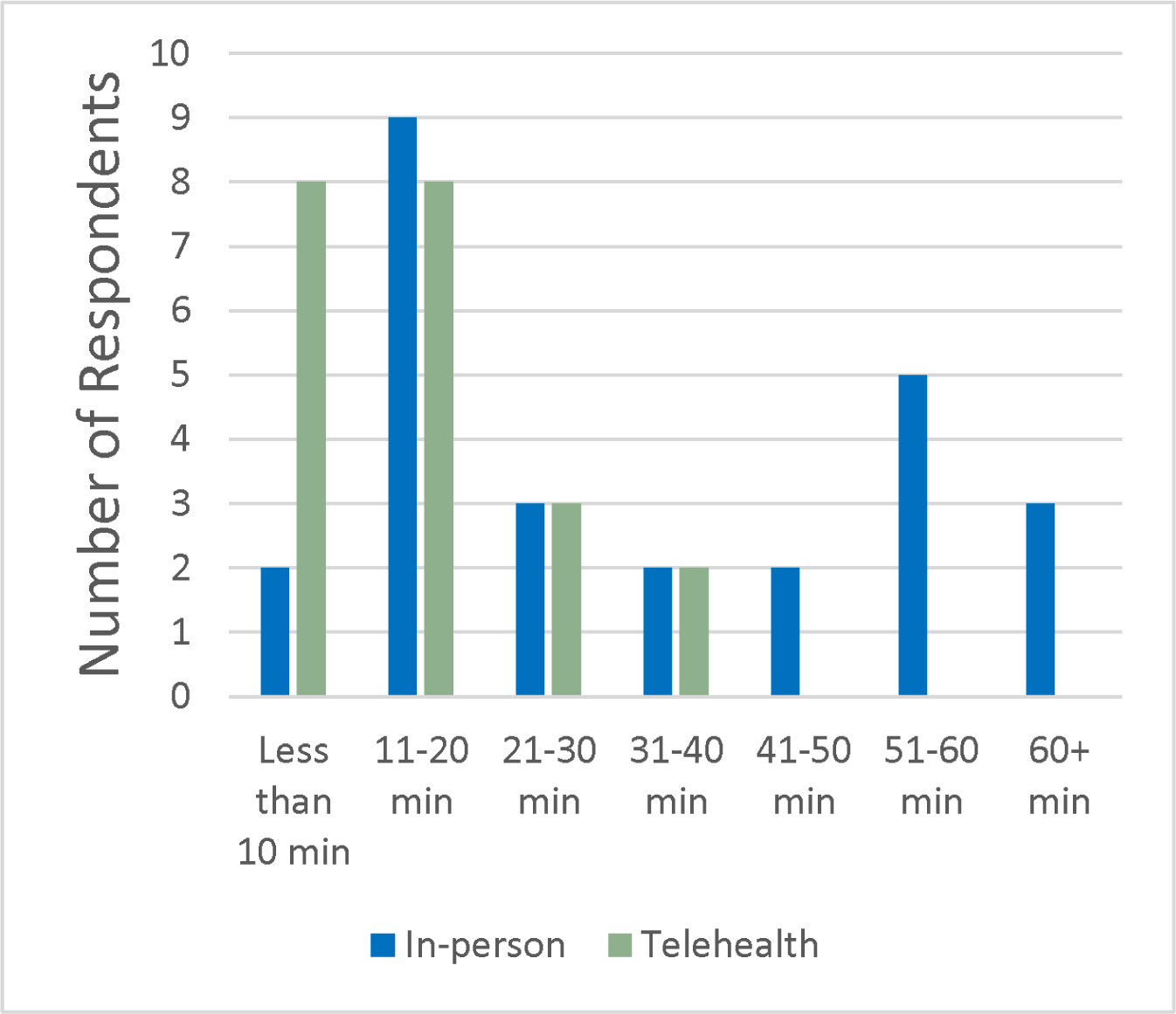
Average Time Preparing for the Session (Including Travel).

**Figure 11.**
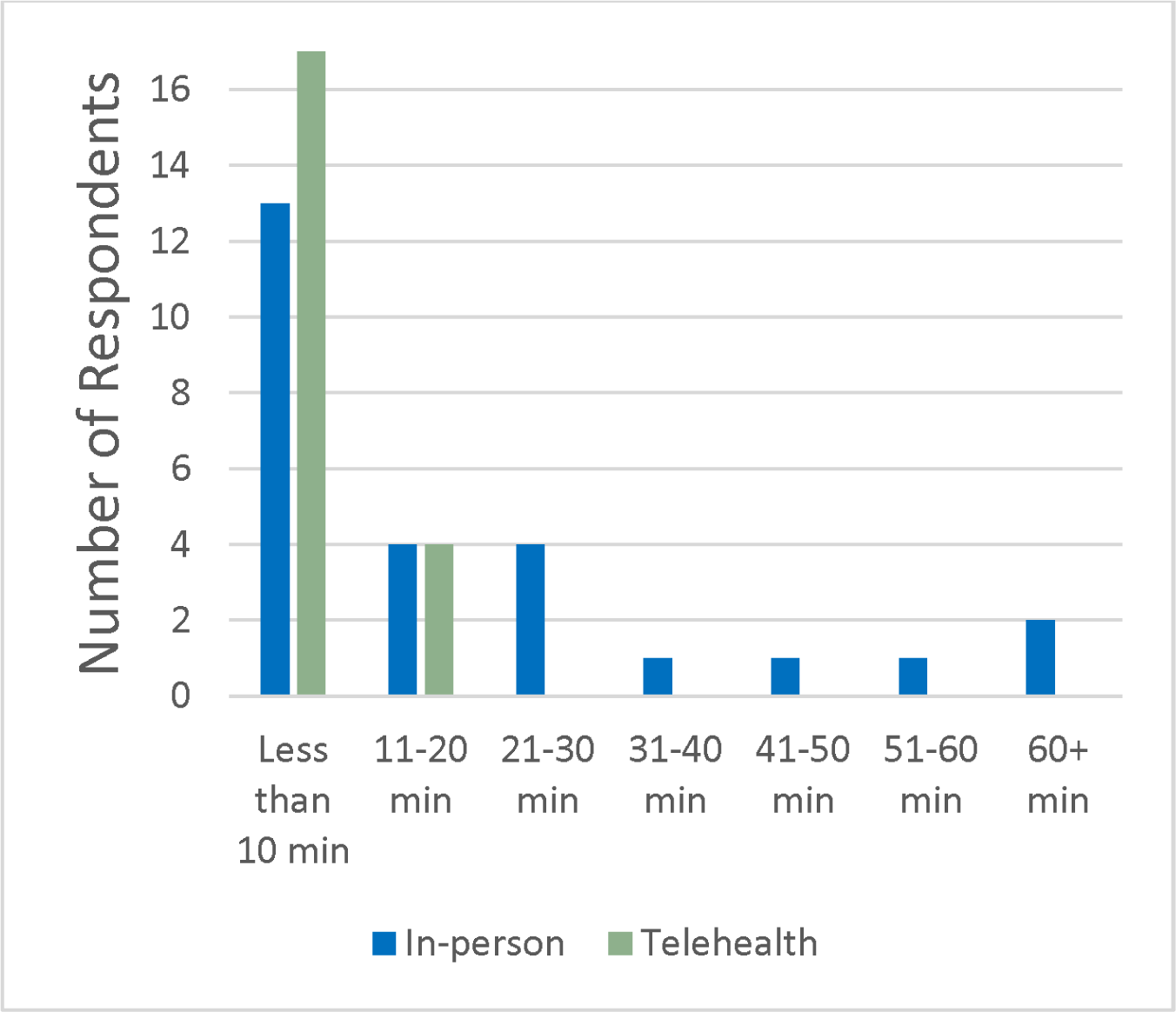
Average Time Transitioning to Normal Activities After Session (Including Travel.)

## DISCUSSION

### Aim 1

*The first aim of this study was to evaluate the efficacy of the SpeechVive device in improving loudness, measured by sound pressure level (SPL), while decreasing pause frequency and duration in an implementation study design focusing on the in-person group*.

To assess this aim, we considered the findings related to the in-person arm of this trial. This study found that the SpeechVive device was effective in improving the loudness of participants, measured by SPL, as implemented in clinical practice. The in-person group increased SPL by +2.45 dB in connected speech, reflecting a perceptible change in loudness. While the effect size was small, the magnitude of change for the in-person group is likely clinically important. Other studies have found similar increases in SPL in controlled designs including one randomized control trial (Richardson, Huber, Kiefer, Kane, et al., 2022; Stathopoulos et al., 2014b). This replication is significant as it shows that these effects are seen in both randomized control studies, as well as in clinical implementation. However, there was no change from pre- to post-treatment without the SpeechVive device on, meaning that the use of the SpeechVive device did not train the participants to speak more loudly. Thus, as we have reported before, we consider the device to have a prosthetic-like effect, like eyeglasses, whereby the device is required for the patient to demonstrate increased loudness (Richardson, Huber, Kiefer, & Snyder, 2022).

The current study found that participants increased their articulation rate pre-to-post treatment. The effect size for this change was moderate; however, the average increase in syllables was small (.09 syllables/second) and likely not clinically significant. This change is very interesting, as we know that a core characteristic of hypokinetic speech is rapid bursts of speech. Interestingly, very few studies have evaluated the impact of behavioral treatment for PD on articulation rate, with no studies specifically evaluating it for the SpeechVive device. The Lombard Effect in typical speakers results in a slowing of rate (Huber et al., 2005; Summers et al., 1988). However, this effect may not be consistently present in people with PD. Sadagopan and Huber (2007) evaluated speech changes when speaking in a noisy environment. They reported that there were no rate changes in the PD group, while older controls reduced their rate of speech in noise. The current study data may support the interpretation that the Lombard effect does not alter rate for people with PD substantially. Further research in this area is necessary to evaluate the replicability of our changes and whether there is a way to use the Lombard effect to slow speech rate in PD.

The device was shown to decrease the number of pauses and total duration of pausing. These findings make sense together. If participants took fewer pauses, this would reduce the total duration of pausing during their speech samples. Mean changes in pause duration were not large enough to be considered clinically significant, but subject-level changes did show high amounts of variability that may be reflective of individual changes. However, changes to the frequency of pauses are likely clinically significant. Other studies have not evaluated the impact of the SpeechVive device on the number of pauses or the duration of pausing. However, some studies have evaluated changes to respiratory kinematics in PD when using the SpeechVive device. If the SpeechVive use resulted in improved respiratory support for speech, participants may need to take fewer pauses to maintain the subglottal pressure necessary for speech. Richardson et al. (2022) and Stathopoulos et al. (2014) both examined respiratory kinematics in people with PD using the SpeechVive device. Richardson et al. (2022) found no significant group-level changes in respiratory patterns. They reported significant participant variation, demonstrating 5 subjects who increased their lung volume initiation with the device on, while 8 subjects decreased their lung volume initiation with the device on. Stathopoulos et al. (2014) found that subjects increased lung volume initiation, termination, and excursion when speaking with the device on as compared to with the device off (Stathopoulos et al., 2014b). The predominant difference between these studies is while Richardson et al. evaluated change over an 8-week treatment duration, Stathopoulos et al. evaluated immediate changes. Further research is needed to examine whether changes in pausing are replicable and whether they do relate to changes in respiratory patterns. Reducing pause frequency and duration are important for conversational dynamics. Increased frequency and duration of pauses can increase the chances of a person with PD losing their conversational turn. The reduction in pause frequency may promote more effective and reciprocal exchanges, improving quality of the person’s conversations. More interactional studies could test these hypotheses.

### Aim 2

*The second aim of this study is to evaluate the effectiveness of the SpeechVive device when administered over teletherapy (telehealth group)*.

Our study found that both groups had statistically similar changes to increasing utterance length, increased articulation rate, decreased number of pauses, and shorter total pause duration. However, there were differences in how SPL was impacted by the device in the telehealth group as compared to the in-person group. While both groups increased their average dB SPL with the device turned on, the participants in the telehealth group demonstrated a smaller increase than the in-person group did (2.45 dB for in-person, .67 dB for telehealth). It is worth noting, however, that both group dB SPL means increased to be within functional limits for older adults (Kent et al., 2023). At baseline, the SPL of the telehealth group was higher (69.87 dB for in-person, 72.40 dB for telehealth), although the difference was not quite statistically significant (p=.08). The difference in baseline SPL may indicate that the participants in the telehealth group were speaking louder because they were talking into the computer, and this may have reduced the Lombard effect on SPL during data collection which occurred over the computer for the telehealth group. One small study has shown that participants increased their loudness on both audio only and video calls compared to normative data (Kisenwether & Anson, 2019). However, this phenomenon has not been well studied. Thus, we may not have measured their “comfortable” intensity for speech during the data collection sessions. There are no data demonstrating how a telehealth interface impacts a person with PD’s performance and whether it might increase awareness of their symptoms or provide an external cue to increase their loudness. Alternatively, the SpeechVive device may not be as effective in increasing loudness when administered via telehealth.

Other studies have found different results when looking at telehealth vs. in-person differences in SPL. Two studies found that patients with PD both in-person and telehealth groups receiving LSVT LOUD therapy increased in their SPL similarly (Constantinescu et al., 2011; Theodoros et al., 2016). A third study reported increased SPL in telehealth after LSVT LOUD treatment in people with PD, but there was no in-person comparison group (Theodoros et al., 2006a). These three studies used controlled research designs, unlike the implementation design for the current study, and the participants were not on their own devices or at home during the prior studies. Further, the three studies had much smaller sample sizes than the current study. Thus, it is difficult to compare the results to the current study, but it is possible that certain therapies are better suited to online therapy than others. More research is needed to better understand the relationship between speech therapy and telehealth services, across a variety of behavioral and device-driven treatments and research designs.

Changes to utterance length differed across the groups as well. People in the telehealth group increased their utterance length (+1 syllable) whereas people in the in-person group decreased utterance length (-.5 syllable). However, neither change was very large nor likely to be clinically significant. Very few studies have evaluated the effect of speech treatment on utterance length in PD, and none of the telehealth studies analyzed this outcome measure. More research to evaluate changes to utterance length resulting from speech therapy, as well as the speech modifications associated with telecommunication, are vital to understanding these results.

In summary, these findings align well with other research that found non-inferiority of telehealth speech and voice treatment for people with PD (Constantinescu et al., 2011; Theodoros et al., 2016). However, because non-inferiority statistics were not assessed in the current study, we cannot say this for certain. More research is needed to confirm and clarify the findings related to SPL in the telehealth group.

### Aim 3

*The third aim of this study is to evaluate if treatment with the SpeechVive device improves patient reported psychosocial and quality of life outcome measures in both the in-person and telehealth modalities*.

In this study, we evaluated psychosocial and quality of life outcome measures. It is of the utmost importance that treatment regimens seek to not only improve qualities of speech and voice, but also to improve a person’s quality of life in terms of their communicative engagement, perceived ability, and overall social and mental health. While this is a vital element of treatment, very few studies have evaluated how speech therapy impacts these outcomes. This study is an important addition to the field in this way.

Our study found that participants in the telehealth group had a 4.5 point mean increase pre- to post-treatment in communicative participation ratings on the CPIB, whereas the in-person group showed a decrease of 1.46 points. These changes may indicate that the device increased communication participation uniquely in the telehealth group. The reasons for this are not well known and more research is vital as this is one of the first studies to use this measure to evaluate communication-related quality of life after treatment.

We did not find significant time (pre-post treatment) or device condition (device on/off) changes for surveys on depression, apathy, ability to participate in social roles and activities, or satisfaction with social roles and activities. We did find a small change satisfaction with social roles and activities in the telehealth group (+1.44) post-treatment. Overall, it is possible that the treatment experience is different for those receiving in-person and telehealth services. More research is needed to better understand these differences and the motivating factors behind them.

This study is unique because it is one of the first to evaluate how speech therapy does or does not modulate changes in these psychosocial outcomes. The change in the CPIB is relevant, as we were unable to find studies that had previously used it to evaluate communication-related quality of life changes. A study by Theodoros et al. (2016) did find communication-related quality of life changes after LSVT LOUD treatment for in-person and telehealth.

Our contributions to the literature regarding other psychosocial changes is additionally important. Our findings for depression contributes to the mixed literature across many fields, suggesting that it is unlikely that rehabilitative treatment plays a unique role in reducing rates of depression for those with PD. However, as this is one of the first studies evaluating changes in speech therapy, it is important to better understand these effects compared to other rehabilitative disciplines that focus on movement-based activities. We were unable to find research supporting therapeutic changes in people with PD for participation and satisfaction with social roles and activities. Our finding that there was an increase in satisfaction with social roles and activities should be investigated with further research to better understand the reasons motivating this improvement, particularly as it was only seen in the telehealth group.

Regarding satisfaction with the device, we found similar levels of satisfaction with the SpeechVive device in both telehealth and in-person groups. Questions regarding ease of use, ability to use the device independently, and that the clinican’s instructions were clear and easy to follow had the highest average scores across all participants. Participants reported unsureness regarding difficulty speaking while using the device, as this question had the lowest average rating across participants, with an average score of 3.5. This study is the first to investigate participant satisfaction for those using the SpeechVive device. Further research is essential to better understand the factors that impact satisfaction and to increase our understanding of participant’s perspectives.

Our study also found that participants were satisfied with telehealth administered therapy. This satisfaction fits well with previous studies that have similarly found high satisfaction with telehealth services for people with PD from a wide variety of disciplines, including speech therapy (Beck et al., 2017c; Dorsey et al., 2016; Griffin et al., 2017; Theodoros et al., 2006b). In addition to the satisfaction with treatment via telehealth, participants in the telehealth group reported less time taken preparing for and transitioning away from speech therapy in their day as compared to the in-person group. Travel to and from therapy can be a significant burden for those with PD as many live in rural areas and as the disease progresses, driving can become more difficult or hazardous (Uc et al., 2017). Thus, this time savings may reflect an important consideration in determining whether to deliver therapy in-person or via telehealth.

### Limitations

One limitation of this study was that it was an implementation design. While implementation science is a vital area of research for clinical treatments, it comes with many natural limitations. The primary limitation is that there was variability across treating SLPs and treatment decisions that they made with the patients. We attempted to mediate this variability by having both SLP and participant as random factors in our statistical analysis. In addition, due to the nature of speech therapy, many SLPs used non-standard treatment protocols, supported by their understanding of the literature and their patient’s needs. This is a vital aspect of person-centered therapy but can increase the amount of variability in implementation studies. However, little treatment was provided to any of the participants outside of using the SpeechVive device. Secondly, we had very few inclusion and exclusion criteria, which increased the variability of our data sample. This was particularly evident regarding wide variety of ages of our participants. Participants in the telehealth group were younger, on average, than those in the in-person group. While both groups had similar speech deficits, we had multiple participants with early-onset PD in our sample. Very little is known about the differences in disease progression for this population which may have impacted our data. The implementation study design also did not allow for randomization since participants were able to select their group. Their choices may have been determined by factors such as wanting to work with a particular SLP, technological comfortability, or accessibility. Another limitation of this study was the loss of data due to technical difficulties. A significant amount of data was lost due to overlapping speech or technological problems. Our study sample was small in comparison to most survey-based research studies, reducing the power of those analyses. A final limitation of this study was the lack of diversity within our sample across multiple factors including race, identity, and socioeconomic status.

### Conclusions

This study demonstrated that the SpeechVive device is effective in improving symptoms associated with hypokinetic dysarthria in PD when utilized in clinical practice. The most significant effects were seen with the device on for the in-person group. This suggests that the device is most effective as an assistive technology rather than a training device. While some changes were seen in the telehealth group, it is likely that this treatment is more effective in in-person settings. More research is needed to better understand how speech changes over telecommunication in both healthy and disordered speakers to support future telehealth studies in speech pathology. More research is also needed to understand how speech treatment, in-person or via telehealth, impacts psychosocial factors such as depression, apathy, and communication participation.

## Data Availability Statement

Deidentified data can be obtained through reasonable request to the corresponding author. A data sharing agreement will be required to obtain the data.

## Data Availability

Deidentified data can be obtained upon reasonable request to Jessica Huber.

## Acknowledgements

Research reported in this publication was supported by the National Institute on Deafness and Other Communication Disorders of the National Institutes of Health under award number R44DC014867. Federal funds in the amount of $277,451 were used to support the larger project from which these data were obtained, covering about 50% of the total costs of the larger project. The content is solely the responsibility of the authors and does not necessarily represent the official views of the National Institutes of Health. We would like to thank the patients with PD and their caregivers who participated in this research.

## APPENDIX A. QUESTIONNAIRES

**Figure A12.**
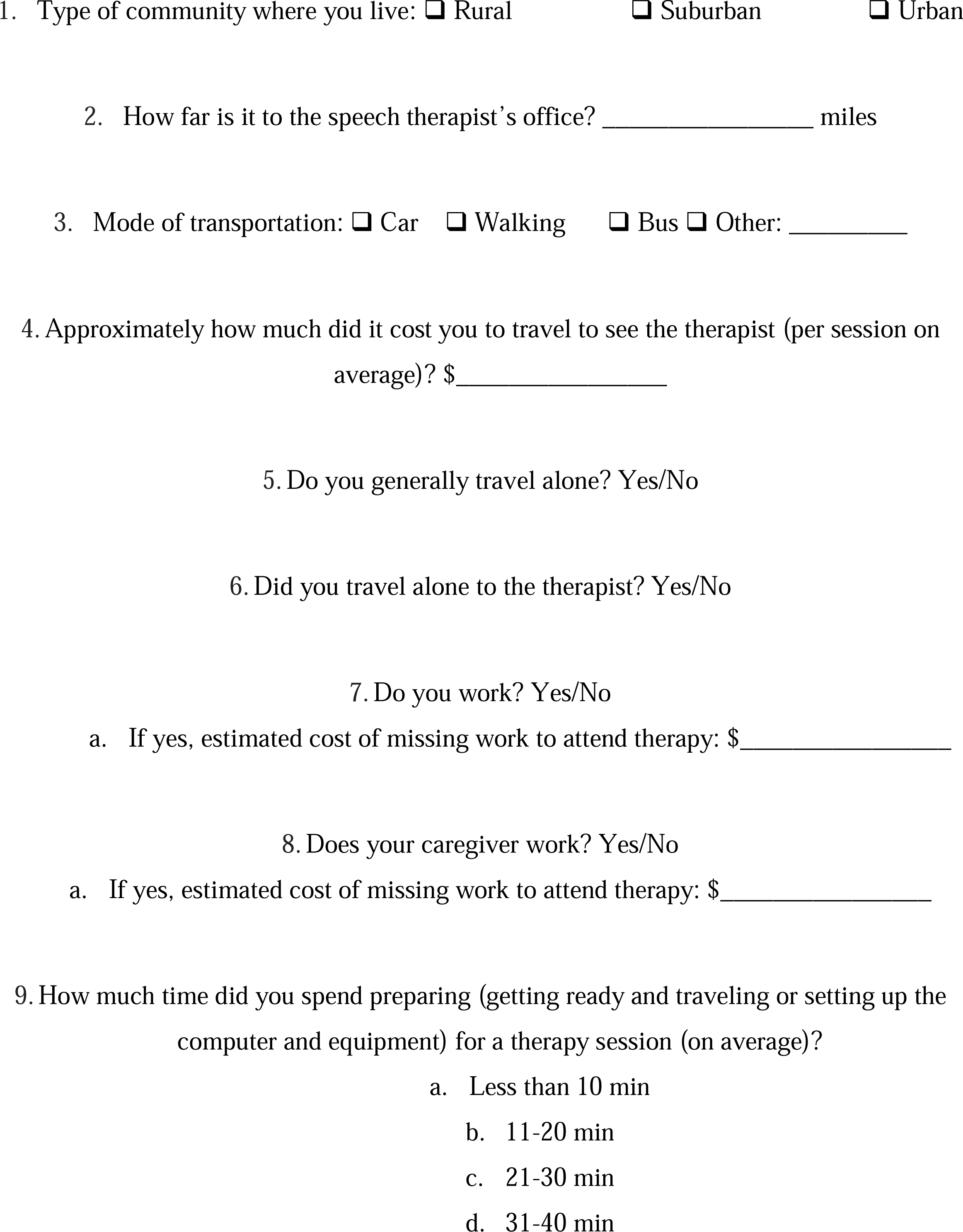
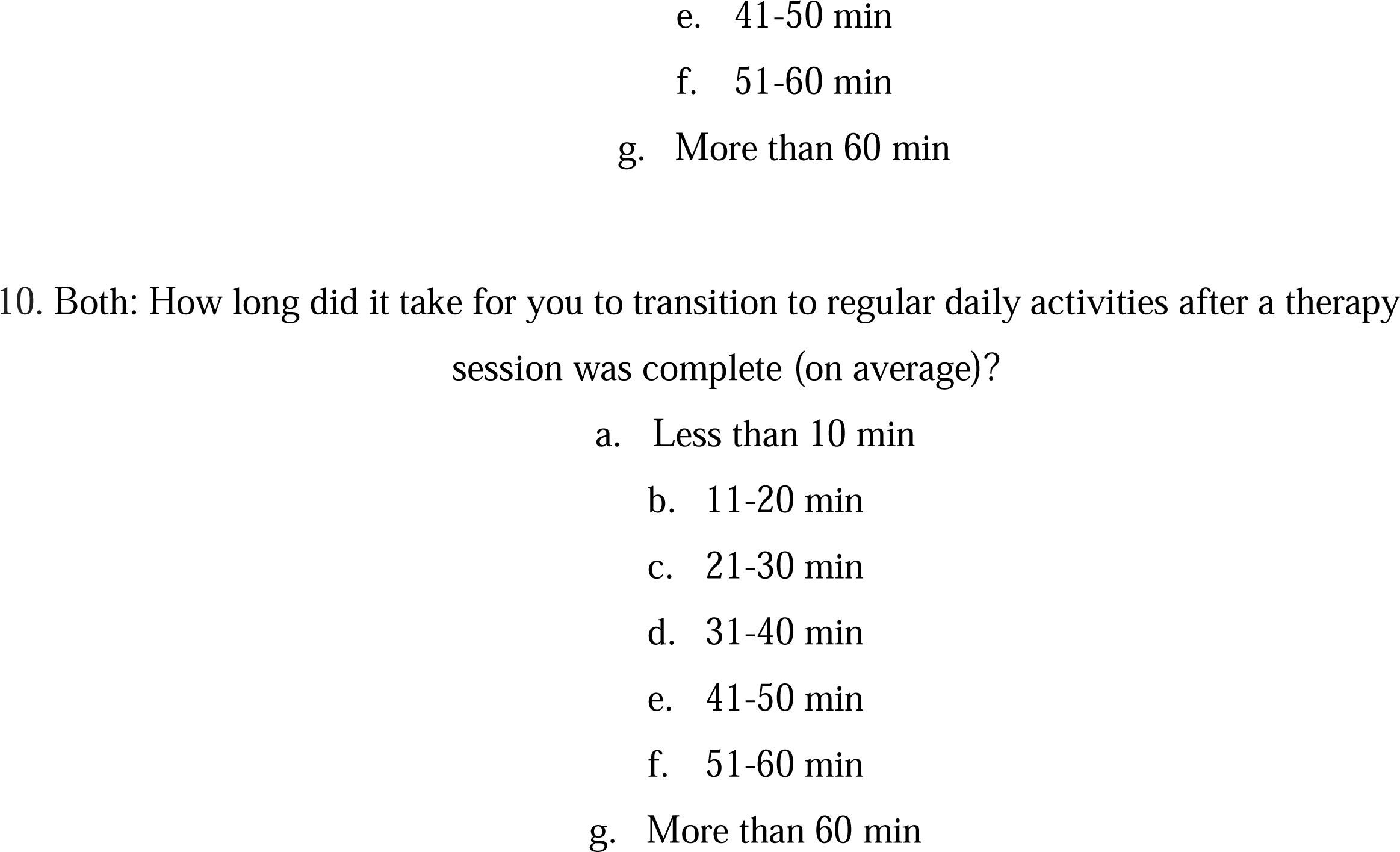
Time/Travel Questionnaire for People with PD and Caregivers.

**Table A8.**
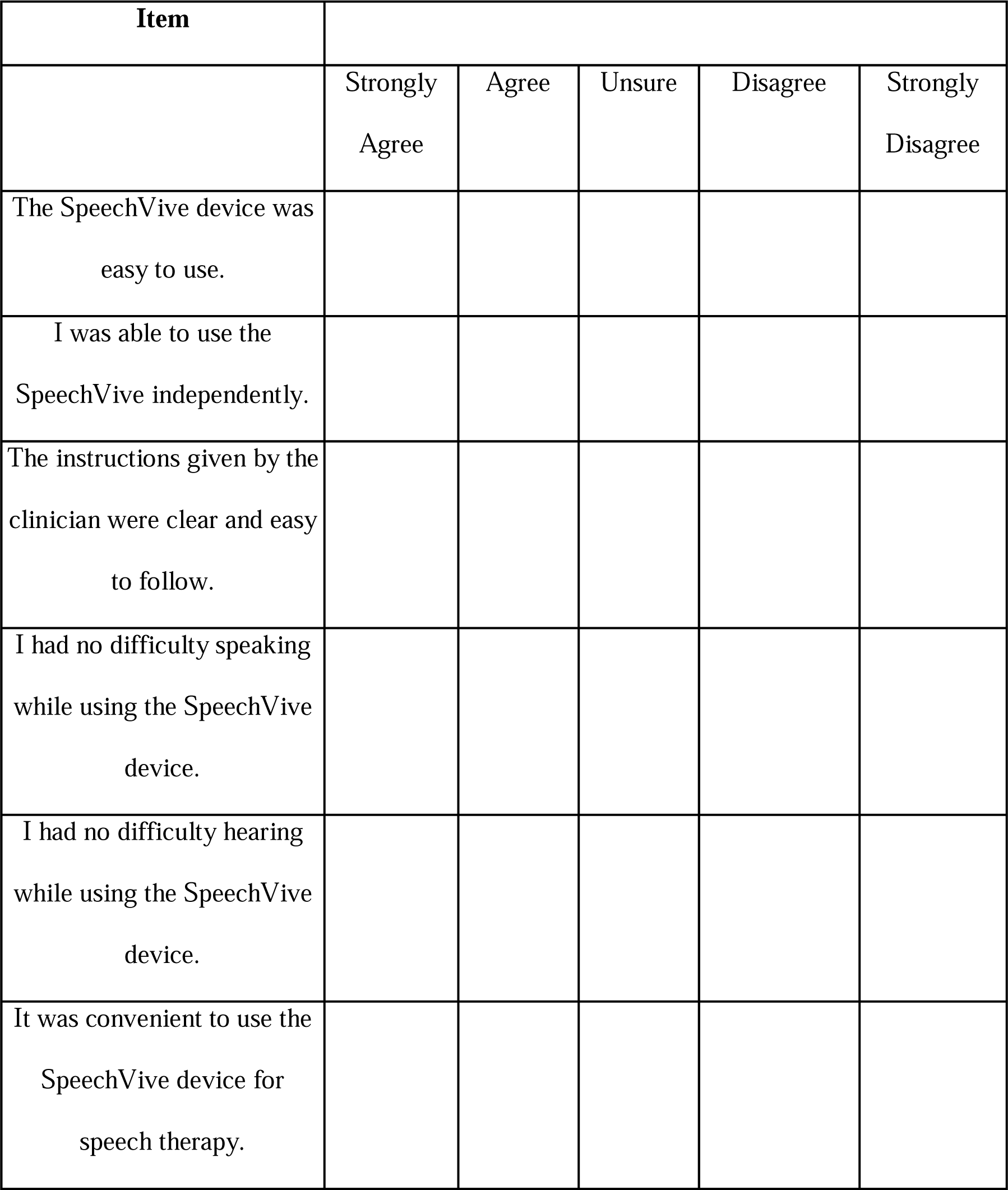
SpeechVive Satisfaction Survey.

**Table A9.**
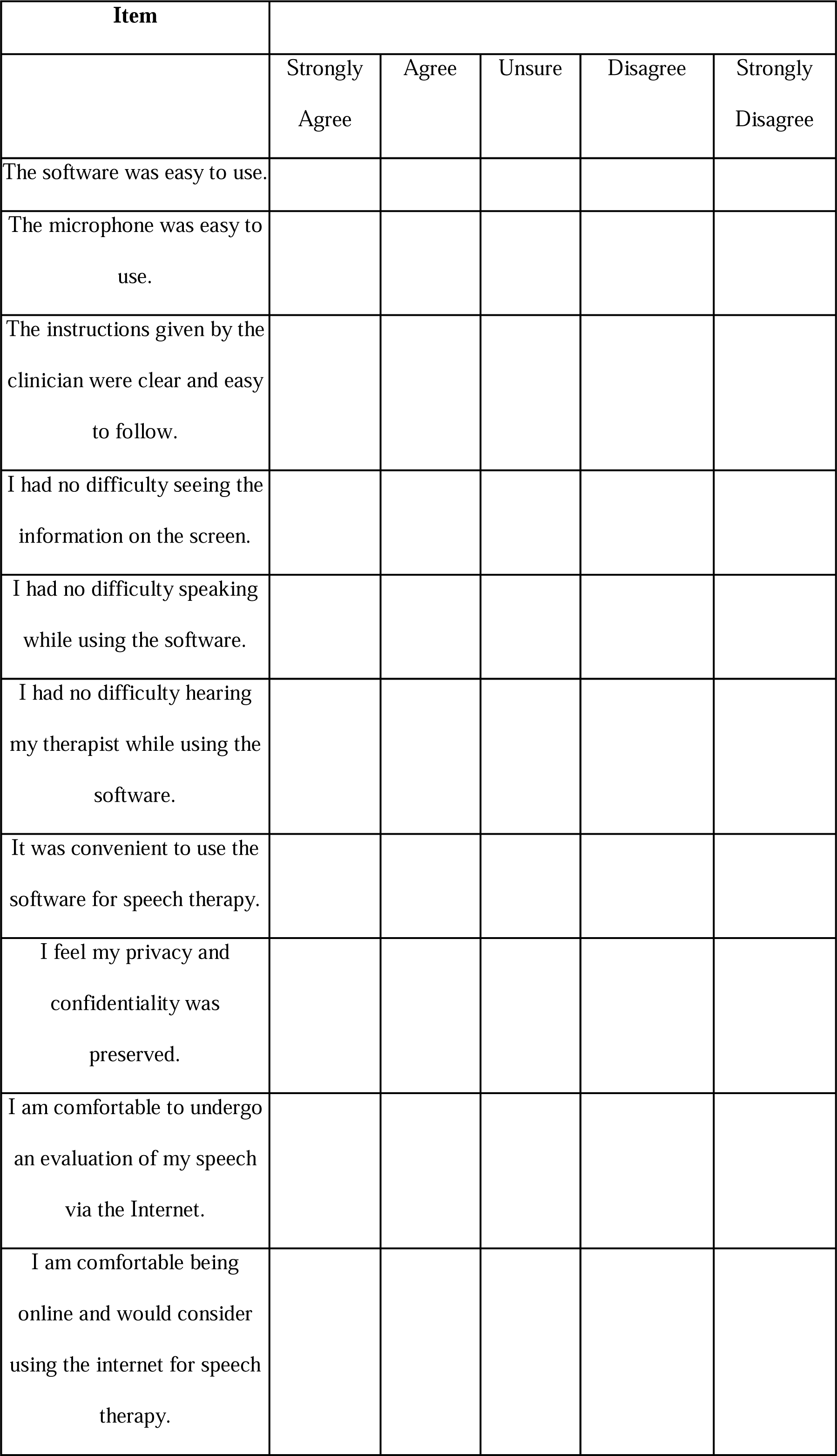
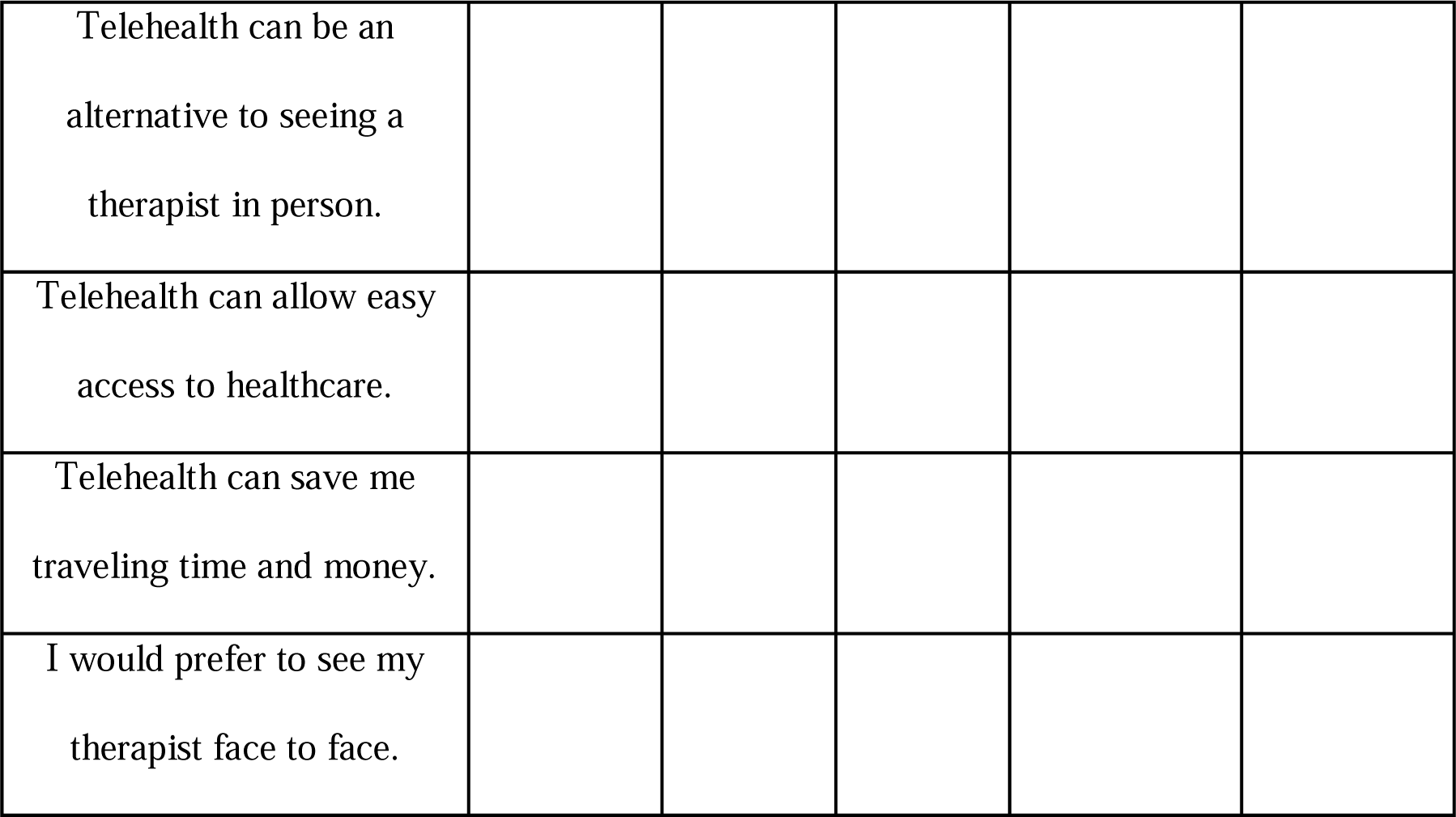
Post Telehealth Satisfaction Survey.

## APPENDIX B. INDIVIDUAL PARTICIPANT DEMOGRAPHICS

**Table B10.**
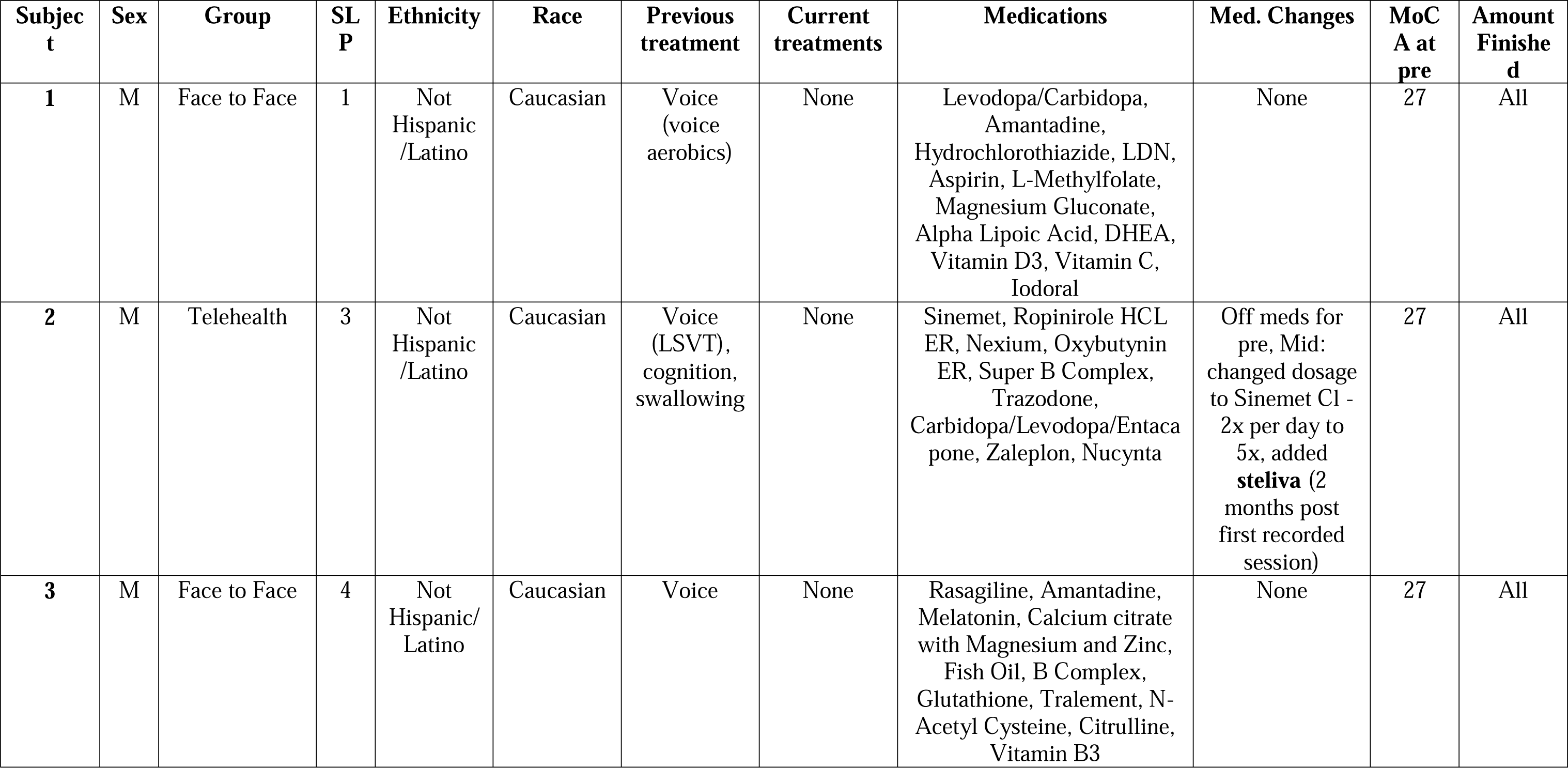
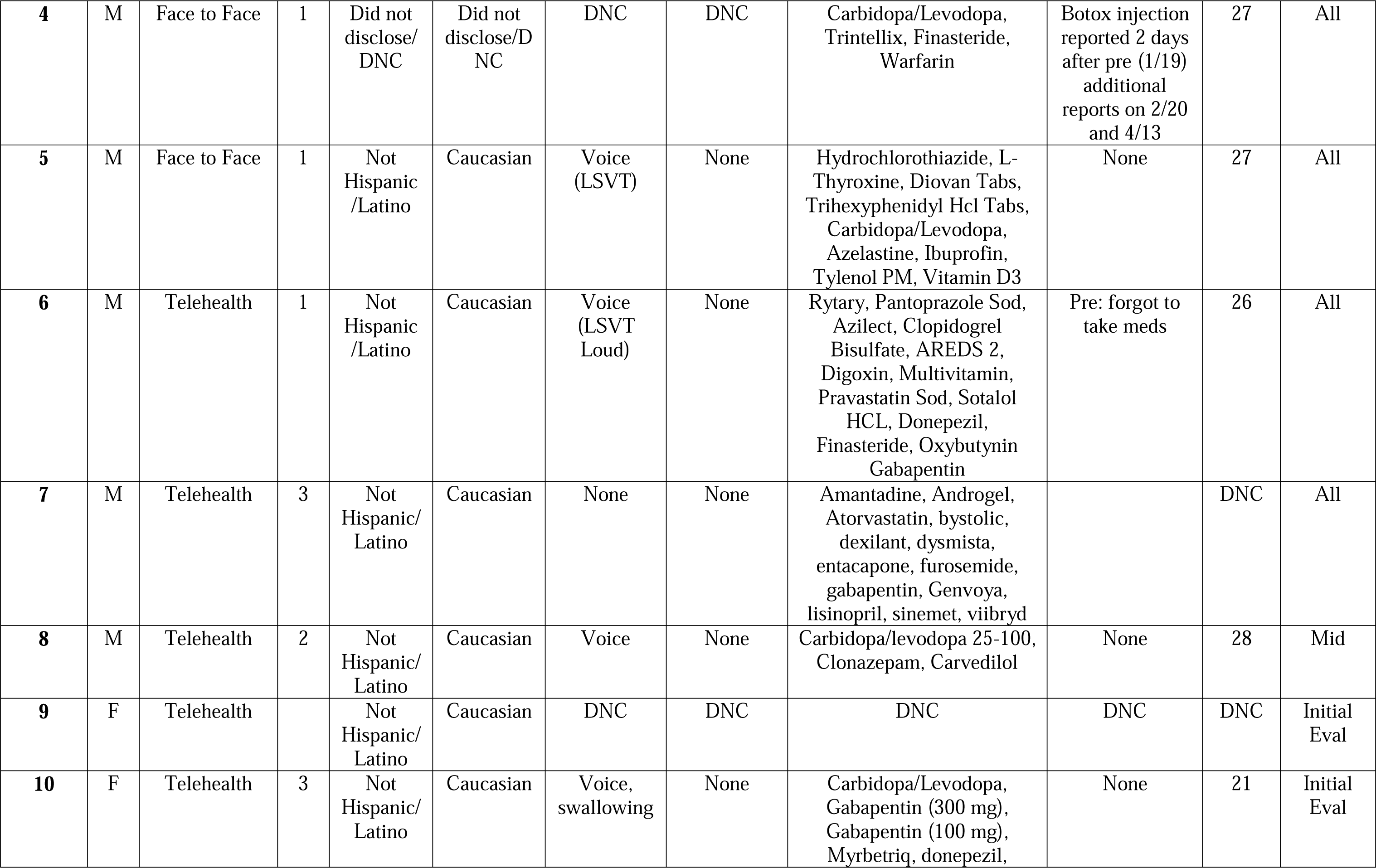
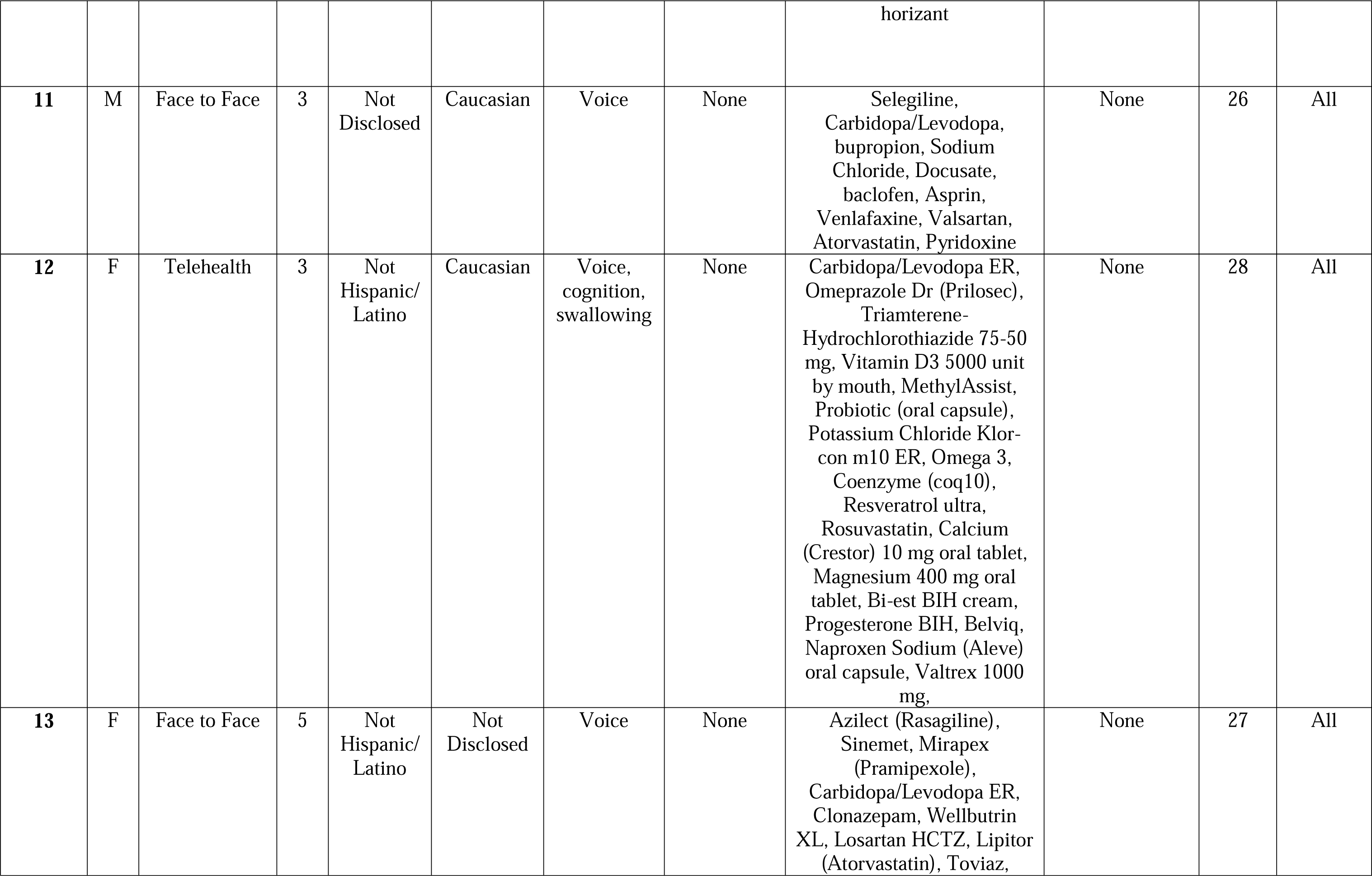
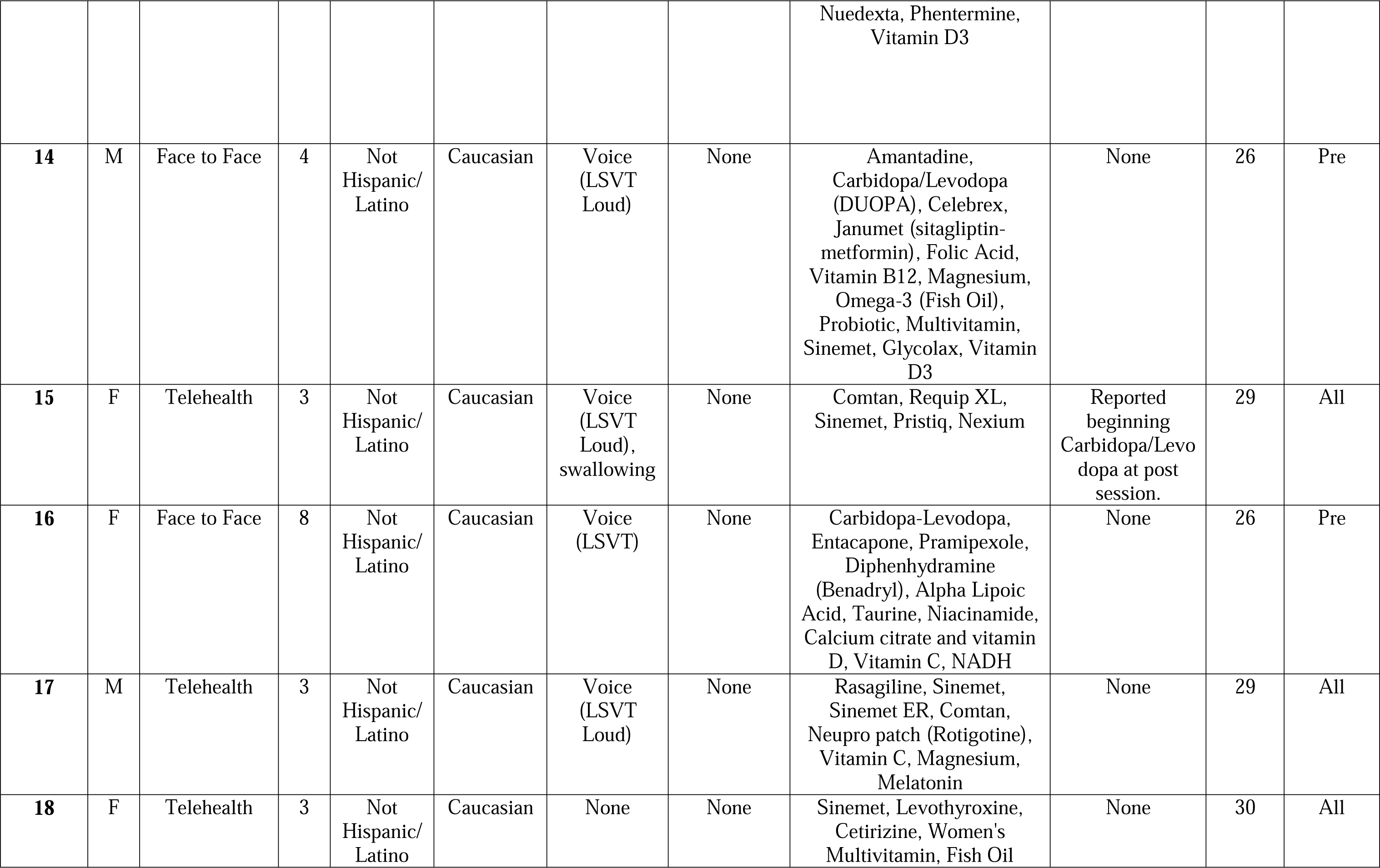
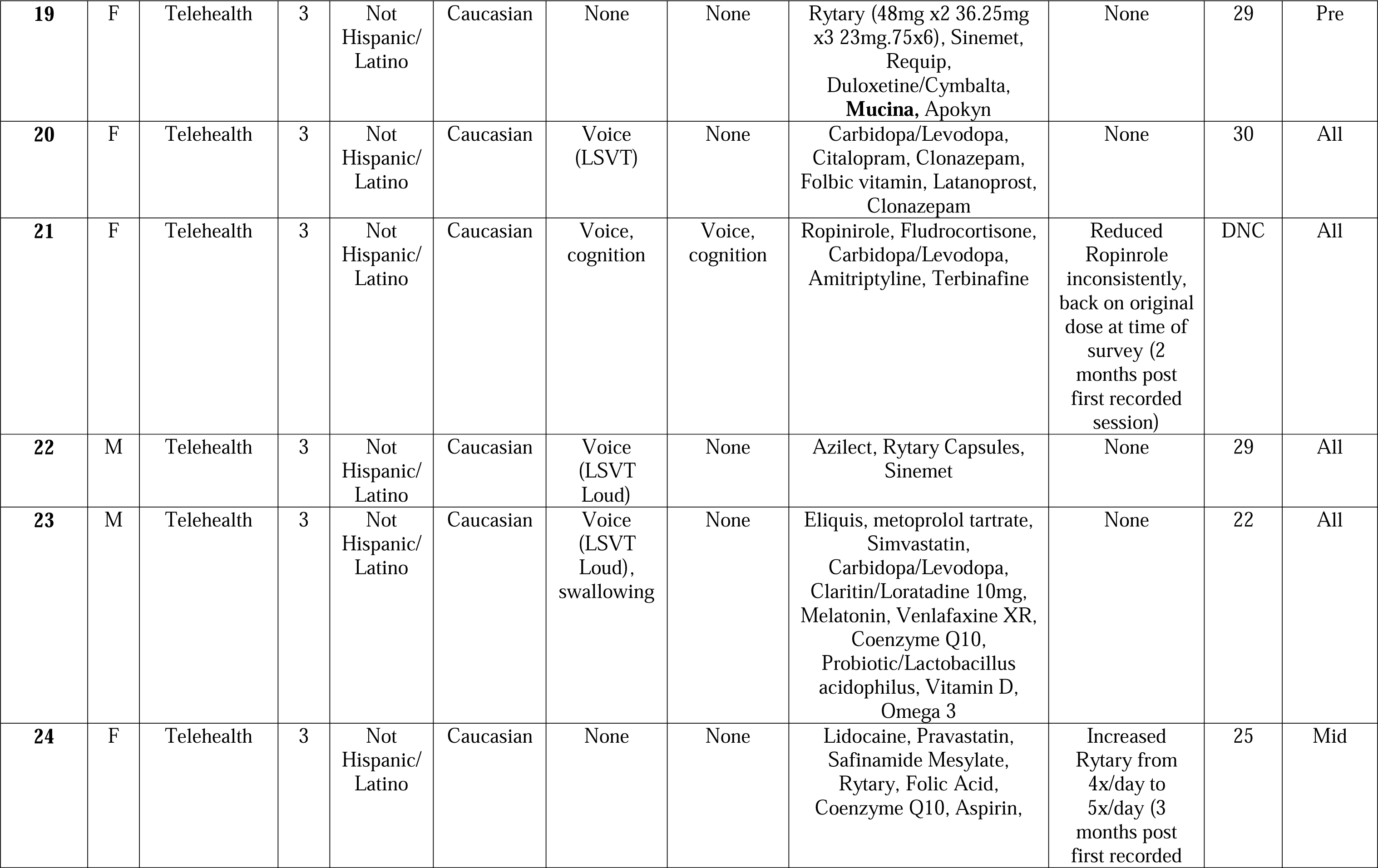
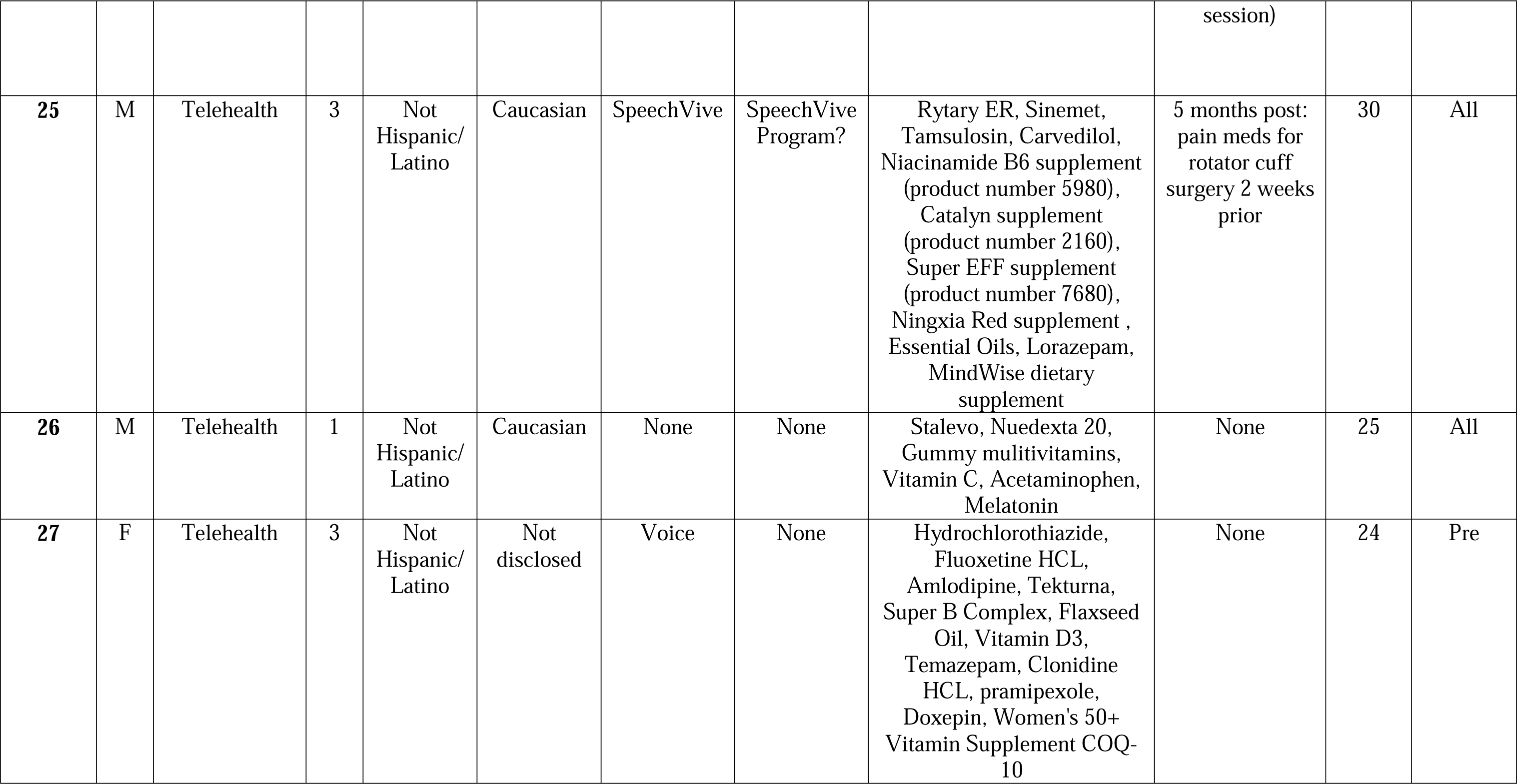
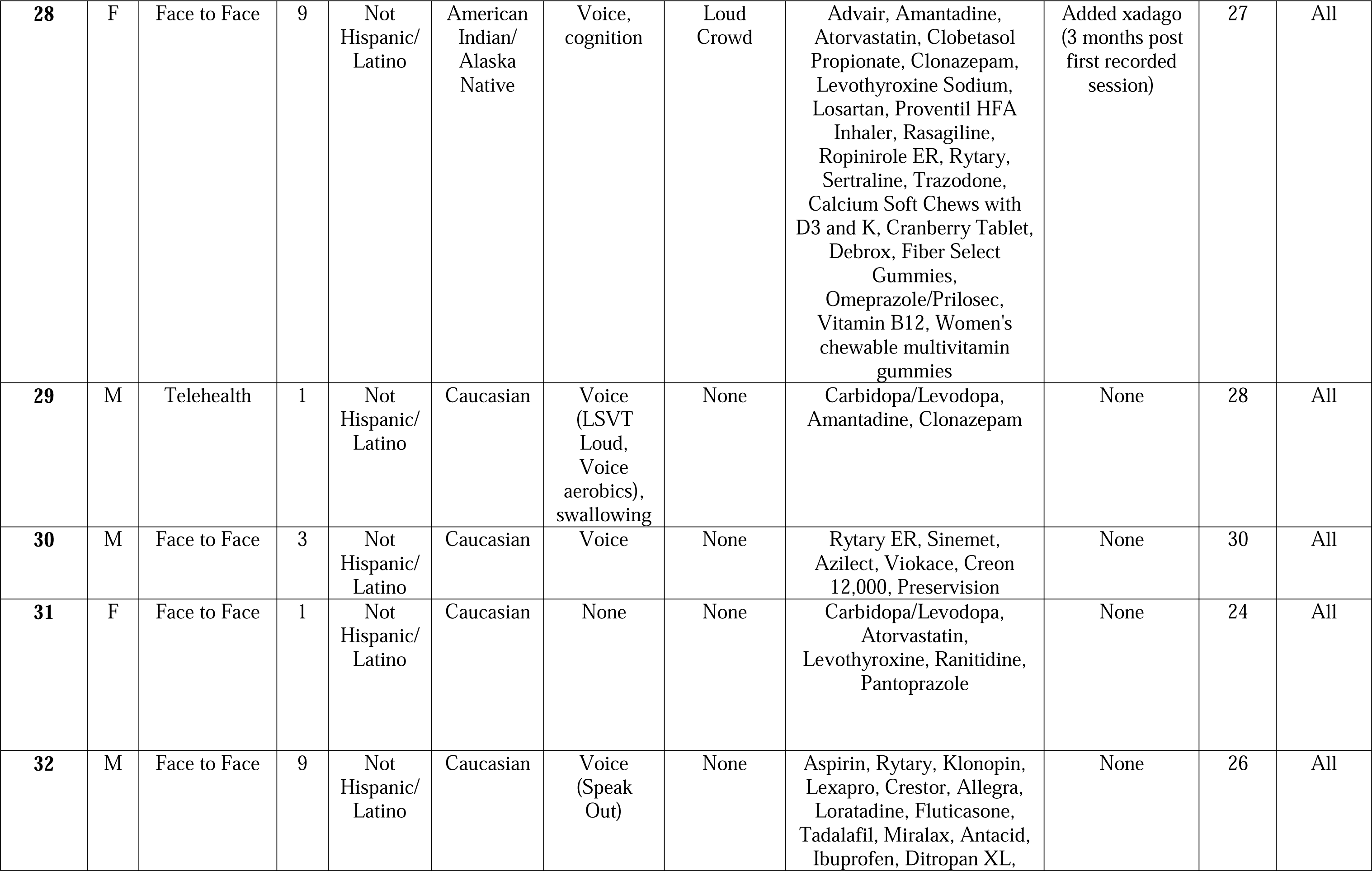
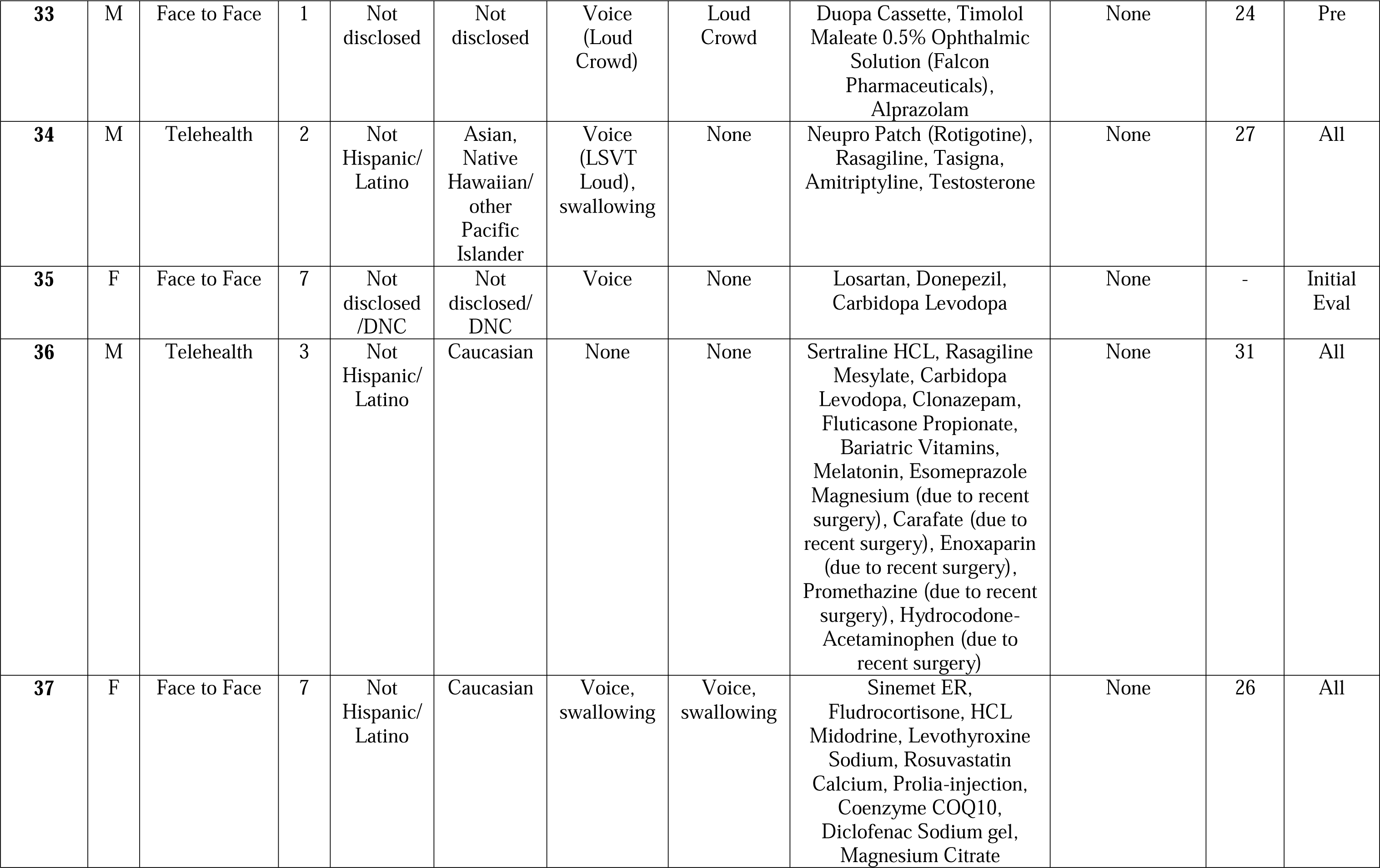
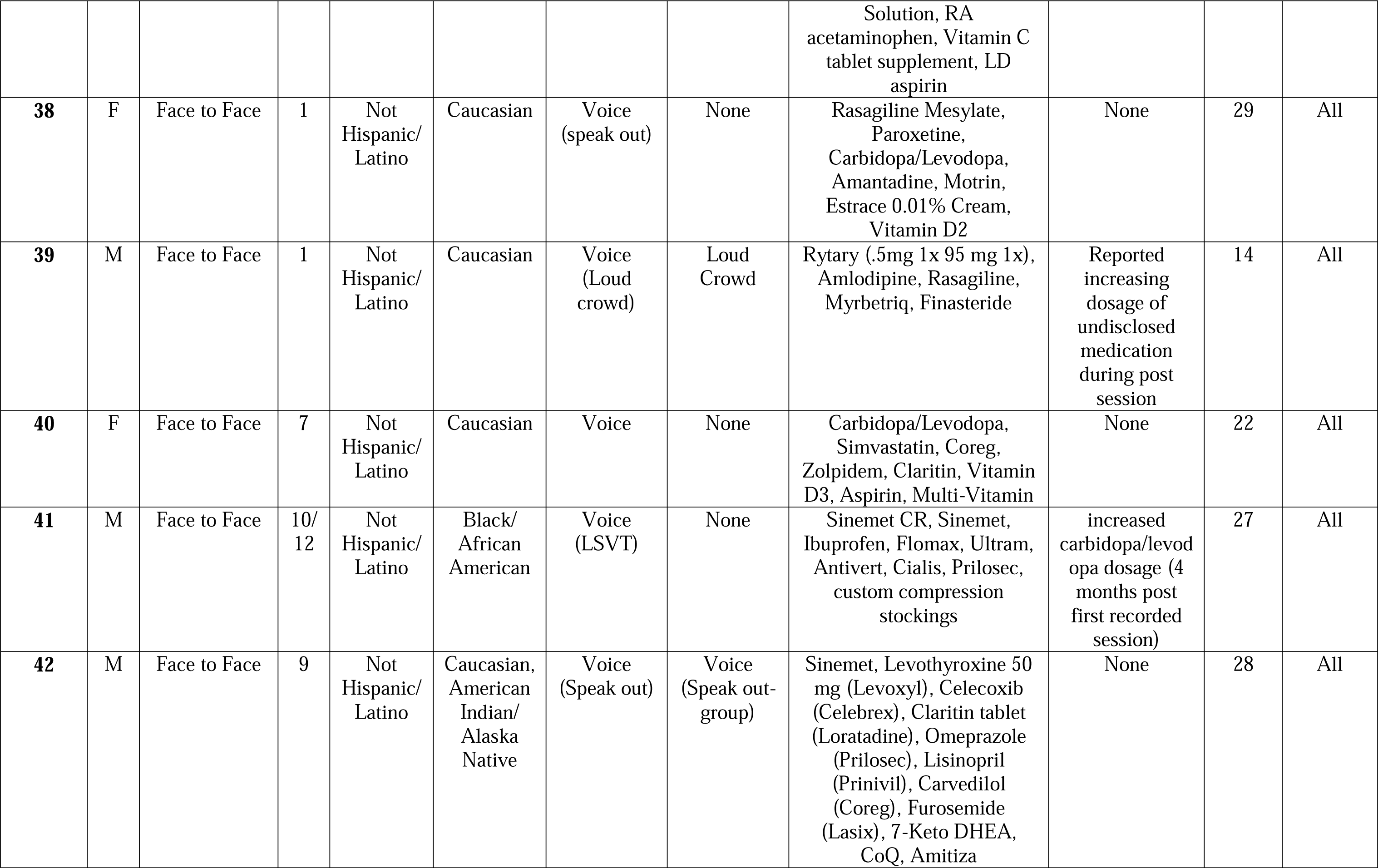
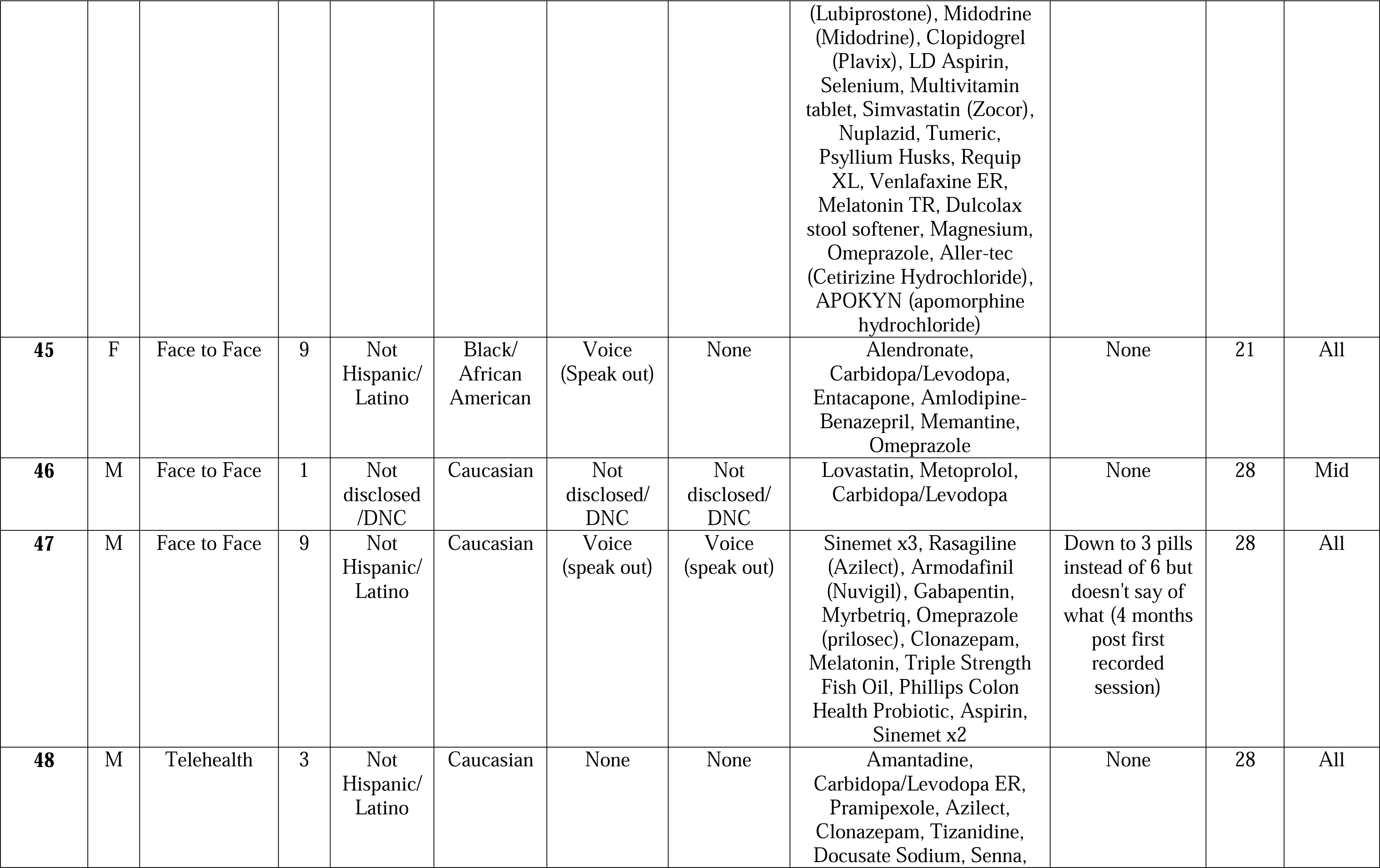
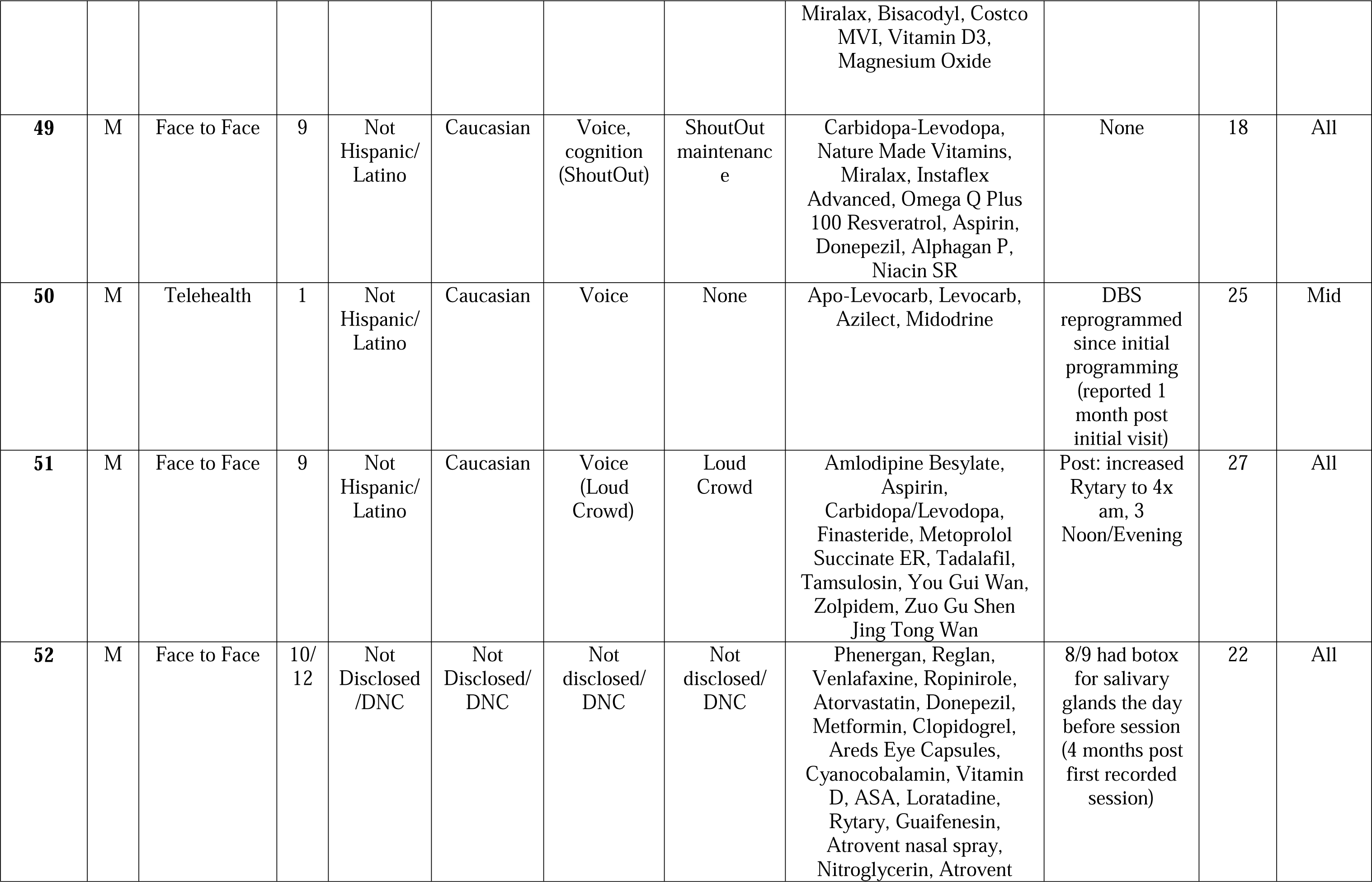
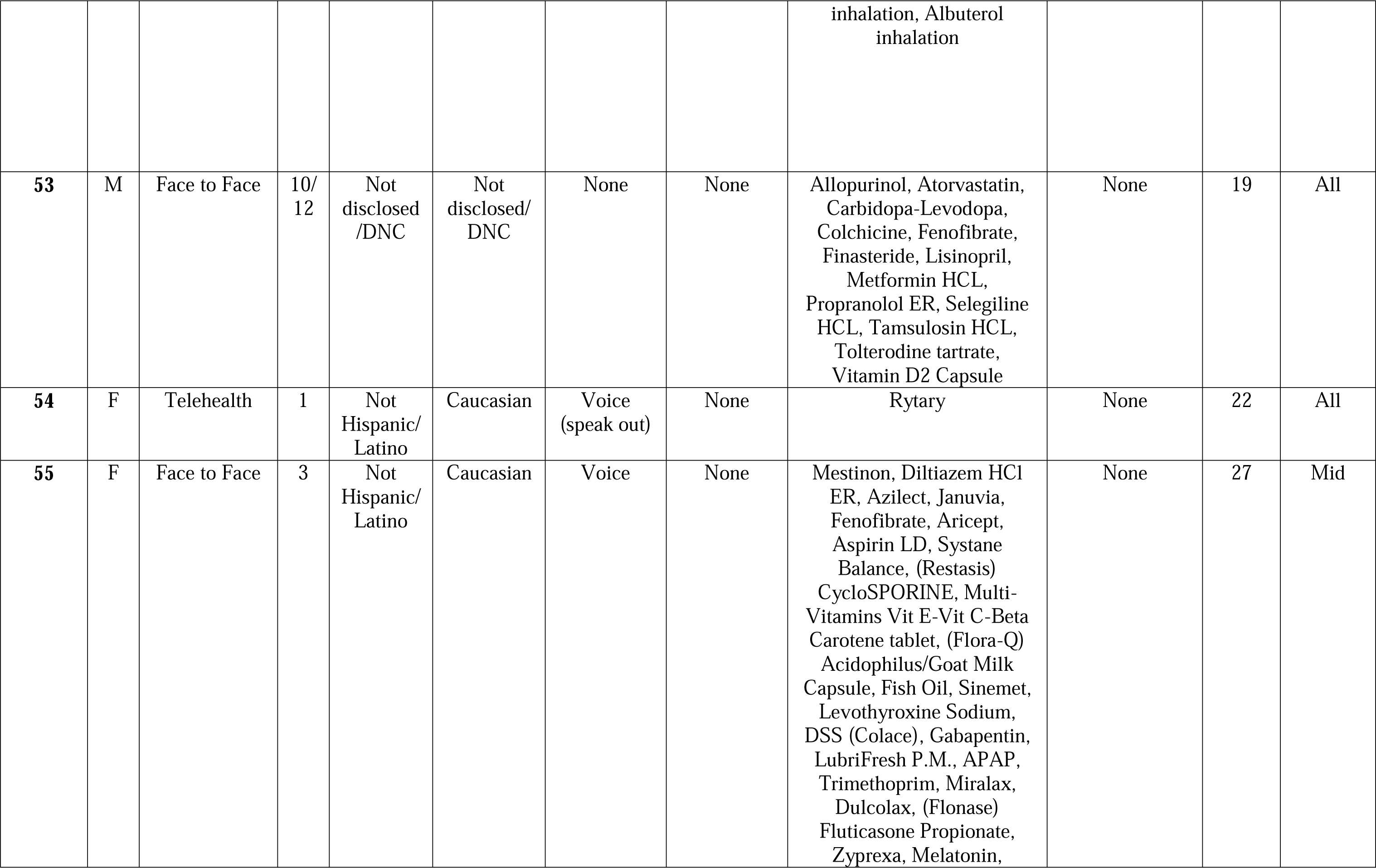
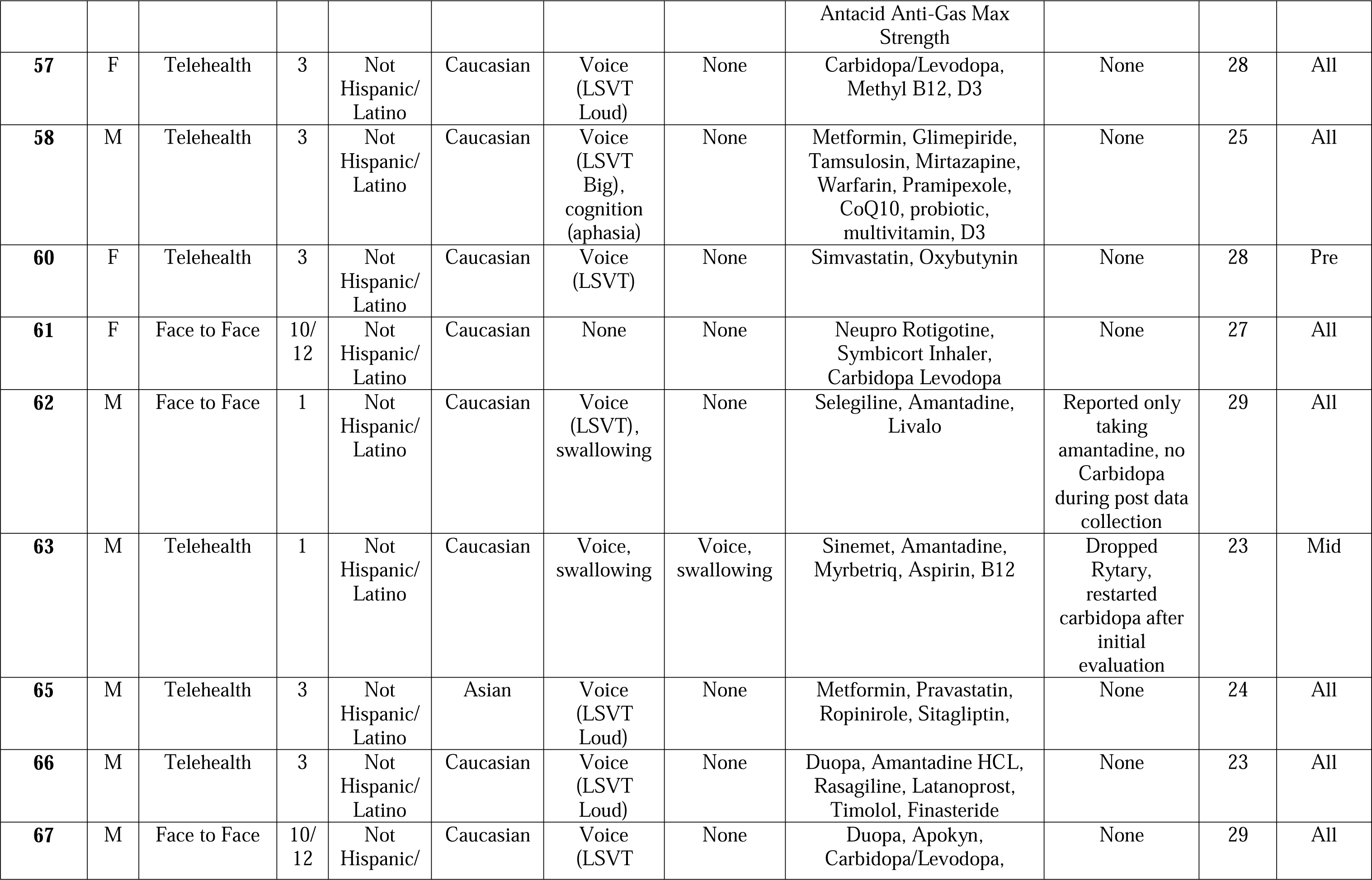
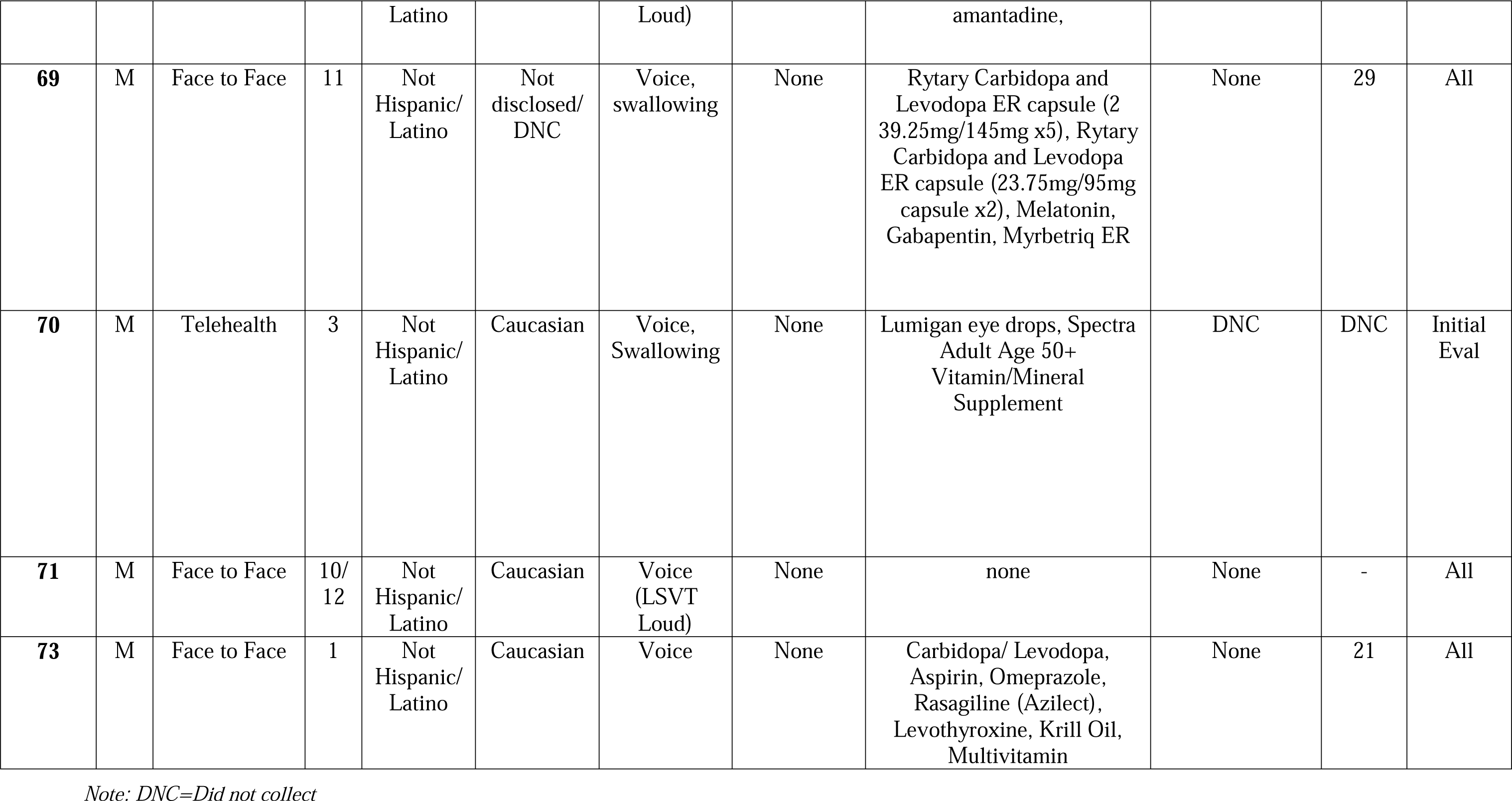
Individual Participant Demographics.

